# Enhanced HIV-1 Control After Antibody Therapy is Associated with Autologous Antibodies and Reservoir Clearance in the RIO trial

**DOI:** 10.1101/2025.11.03.25339415

**Authors:** Marcilio J. Fumagalli, Anna Kaczynska, Marius Allombert, Ágata Lopes-Ribeiro, Mariângela de Oliveira Silva, Brianna J. Hernandez, Christy Lavine, Sebastian M. Espinosa, Helen Brown, Hanna Box, Emanuela Falaschetti, Louise-Rae Cherrill, Tamara Elliott, Ming Lee, Julie Fox, Sarah Pett, Amanda Clarke, Alison Uriel, Ole Sogaard, Rebecca Sutherland, Marta Boffito, Sabine Kinloch-Loes, Chloe Orkin, Simon Collins, Panagiota Zacharopoulou, Nicola Robison, Cintia Bittar, Thiago Oliveira, Anna Gazumyan, Mila Jankovic, Michael S. Seaman, John Frater, Marina Caskey, Sarah Fidler, Michel C. Nussenzweig

**Affiliations:** Laboratory of Molecular Immunology, The Rockefeller University, New York, NY USA; Humoral Immunology Unit, Institute Pasteur, Université Paris Cité, Paris, France; Laboratory of Basic and Applied Virology, Microbiology department, Institute of Biological Sciences, Federal University of Minas Gerais, Belo Horizonte, MG, Brazil; Laboratory of Antigen Targeting to Dendritic Cells, Parasitology Department, University of São Paulo, São Paulo, SP, Brazil; Center for Virology and Vaccine Research, Beth Israel Deaconess Medical Center, Harvard Medical School, Boston, MA, USA; Nuffield Department of Medicine, University of Oxford, Oxford, UK and the Oxford NIHR Biomedical Research Centre, Oxford, UK; School of Public Health, Imperial Clinical Trials Unit, Imperial College London, London, UK; NIHR Imperial Biomedical Research Centre, Imperial College London, London, UK; Department of Infectious Disease, Imperial College London, London, UK; Guy’s and St Thomas Hospital NHS Foundation Trust, London, UK; University College London, London, UK; Central and Northwest London NHS Foundation Trust, London, UK; University Hospitals Sussex NHS Foundation Trust, Brighton, UK; Brighton & Sussex Medical School, Brighton, UK; Manchester University NHS Foundation Trust, Manchester, UK; Department of Clinical Medicine, Aarhus University Hospital, Aarhus, Denmark; Western General Hospital, NHS Lothian Trust, Edinburgh, UK; Chelsea and Westminster Hospital, London, UK; Royal Free London NHS Foundation Trust, London, UK; Queen Mary University of London, London, UK; Barts Health NHS Trust, London, UK; HIV i-Base, London, UK; Department of Genetics, Ribeirao Preto Medical School, University of Sao Paulo, Ribeirão Preto, SP, Brazil; NIHR Oxford Biomedical Research Centre, University of Oxford, Oxford, UK; Howard Hughes Medical Institute, The Rockefeller University, New York, NY, USA

## Abstract

RIO is an ongoing double blind randomized placebo controlled human trial in which participants that started antiretroviral therapy (ART) during primary or early-stage infection underwent treatment interruption and were randomly assigned to a group receiving one or two doses of two long acting broadly neutralizing antibodies 3BNC117-LS and 10-1074-LS (Arm A, n = 34), or saline (Arm B, n = 34). The primary clinical study met its primary endpoint and was published elsewhere, demonstrating significantly delayed viral rebound and prolonged ART-free control in participants receiving bNAbs compared to placebo. The infusions were generally safe and well tolerated. Here we report on pre-specified secondary analyses examining viral rebound, antibody sensitivity, and reservoir dynamics, as well as exploratory analyses of rebound virus evolution and autologous antibody activity. Pre-infusion reservoir measurements showed low levels of intact proviral HIV-1 DNA in circulating CD4+ T cells in both arms. The rebounding viruses in participants that received the antibodies, showed significant selection for resistance to 10-1074-LS but not to 3BNC117-LS. Notably, there was a significant correlation between greater initial reservoir sensitivity to autologous antibodies and to 10-1074 and time to rebound. Finally, comparison of pre-infusion and pre-rebound HIV-1 proviral reservoir showed a decrease in intact but not defective proviruses in Arm A but not in Arm B. Together, these findings suggest that baseline humoral immunity and prolonged exposure to LS-bNAbs may shape post-treatment viral rebound dynamics in individuals that initiate ART during primary infection.

ClinicalTrials.gov identifier: NCT04319367

## Introduction

Upon entry into CD4+ T cells the HIV-1 RNA genome is reverse transcribed and preferentially integrated into transcriptionally active sites in the host genome ^1^. High level transcription of replication-competent proviruses leads to cell death and, if untreated, almost invariably results in lethal immunodeficiency ^2^. Anti-retroviral therapy (ART) prevents the spread of infection and immunodeficiency but fails to eliminate a reservoir of latent proviruses found primarily in long-lived, resting memory CD4+ T cells ^3–6^. These reservoir cells express viral RNA at levels below the threshold required to either induce cytopathic effects or elicit robust immune-mediated clearance, yet they can retain the capacity to reinitiate systemic infection upon ART interruption^7–9^.

The HIV-1 reservoir contains both intact and defective proviruses and has been characterized by several different methods including viral outgrowth assays, digital droplet polymerase chain reaction (ddPCR), and whole genome amplification and sequencing (Q4PCR) ^10–13^. Whereas the size of the defective reservoir is relatively stable over time on ART, the intact reservoir shows early rapid decay which slows to a half-life of approximately 4.5 years, with possible further prolongation after 7 years on therapy ^14–17^. Intact proviruses remaining in the reservoir after long-term therapy tend to reside in areas of the genome that are less transcriptionally active ^18–20^. Although the mechanism of selection against the reservoir viruses integrated into transcriptionally active parts of the genome is not precisely understood; selection is thought to be mediated by a combination of HIV-1 cytopathic effects and host immune responses.

Passively administered antibodies can enhance host immune responses and induce simian HIV control by a CD8^+^ T cell-dependent mechanism ^21–23^. In addition, several small studies in humans have reported that broadly neutralizing antibody (bNAb) therapy is associated with accelerated clearance of infected cells, delayed viral rebound and a higher-than-expected rate of prolonged control ^24–29^. Control has been associated with increased HIV-1 specific CD8^+^ T cell responses in some but not all studies ^29–32^. In addition, observational studies in post treatment controllers, people living with HIV (PLWH), and tissue culture experiments suggest that autologous antibodies may also put pressure on the reservoir ^33–36^.

The RIO trial is a randomized, double-blind, placebo-controlled phase II study evaluating the long-acting bNAbs 3BNC117-LS and 10-1074-LS administered during analytical treatment interruption (ATI) in individuals who initiated ART during primary or early HIV infection ^37,38^. Here, we report on pre-specified secondary and exploratory analyses including viral reservoir composition and antibody sensitivity that shape rebound dynamics and post-treatment control in the RIO trial.

## Results

### Trial overview

The RIO trial enrolled 68 participants that were randomized 1:1 to receive two bNAbs, 3BNC117-LS and 10-1074-LS (Arm A), or placebo (Arm B) 2 days before ATI. Baseline demographic and clinical characteristics were balanced between the two study arms (Supplementary Table 1). The primary outcomes, reported elsewhere ^37^, were safety and time to viral rebound 20 weeks after ATI. The infusions were generally safe. ART was restarted if the viral load was >100,000 HIV-1 copies ml^-1^ for 2 consecutive weeks, or > 1,000 HIV-1 copies ml^-1^ for 6 weeks or CD4+ T cell count <350 cells ml^-1^, or clinical symptoms or participant preference^37^. Early rebound before week 20 occurred in 8 Arm A and 30 Arm B participants: 75% versus 11% (P < 0.001, Extended Data Fig. 1). The median time to ART restart was 45.4 weeks in Arm A, where 7 of 29 participants (24%) remained off ART at 96 weeks, compared to 4.6 weeks in Arm B, where only 2 of 32 (6%) remained undetectable throughout 96 weeks of observation (P < 0.001, Extended Data Fig. 1 and Supplementary Table 1) ^37^. Notably, 13 of the 29 individuals in Arm A restarted ART before meeting virological restart criteria ^37^.

Prespecified secondary outcomes documented here include quantitation of viral reservoir size and dynamics, and reservoir viral sensitivity to bNAbs and autologous antibodies. Prespecified exploratory outcomes include rebound virus sequencing, phylogenetic relationships, antibody resistance patterns, and antibody correlates of viral control. Additional post hoc analyses were performed to evaluate the relationship between reservoir size, viral sensitivity, and clinical outcomes.

### Baseline reservoir

To examine the HIV-1 proviral reservoir in individuals enrolled in the RIO trial, we assayed pre-intervention peripheral blood mononuclear cells. Reservoir size was estimated by ddPCR and Q4PCR ^10,11^. ddPCR documents the presence of 2 conserved regions in the provirus, the packaging signal and envelope ^10^. It is more sensitive, less expensive and scalable than Q4PCR^11^. However, it does not allow sequence confirmation and therefore some of the proviruses that are assigned as intact are in fact defective, a problem that can increase with time because the intact reservoir decays faster than the defective ^39^. Q4PCR provides sequence information and direct confirmation of whether a provirus is intact or defective, but it is less sensitive than ddPCR because it requires amplification of the 9 kilobase proviral genome ^40^.

To examine the pre-intervention reservoir in the RIO participants we performed both assays. Individuals in the 2 arms (ddPCR: Arm A n=32, Arm B n=29, Q4PCR: Arm A n = 30, Arm B n= 28) showed similar numbers of intact and defective proviruses in ddPCR and Q4PCR assays (Extended Data Fig. 2a and b, and Supplementary Table 2). The geometric mean number of intact proviruses per million CD4+ T cells was 8.58 and 6.94 by ddPCR (p=0.5), and 0.27 and 0.28 by Q4PCR (p=0.5), for Arms A and B, respectively (Extended Data Fig. 2a). The relatively small size of the reservoir in this cohort is consistent with early therapy ^41–43^.

We were able to recover 141 and 91 intact proviral reservoir sequences from baseline samples from 15 and 11 individuals in Arms A and B, respectively (Fig. 1a and b, Supplementary Table 3). We were unable to recover intact proviruses from the remaining participants due to their small reservoirs. As expected, each participant harbored a separate non-overlapping group of intact HIV-1 proviral sequences (Fig. 1a and b). Despite early therapy and the small reservoir sizes, intact proviruses showed a median level of clonality of 57% when considering all individuals from whom we were able to obtain 3 or more intact proviral sequences (n =10 for Arm A and n = 8 for Arm B) ^12,44–46^ (Fig. 1c and Supplementary Table 3).

**Figure 1.**
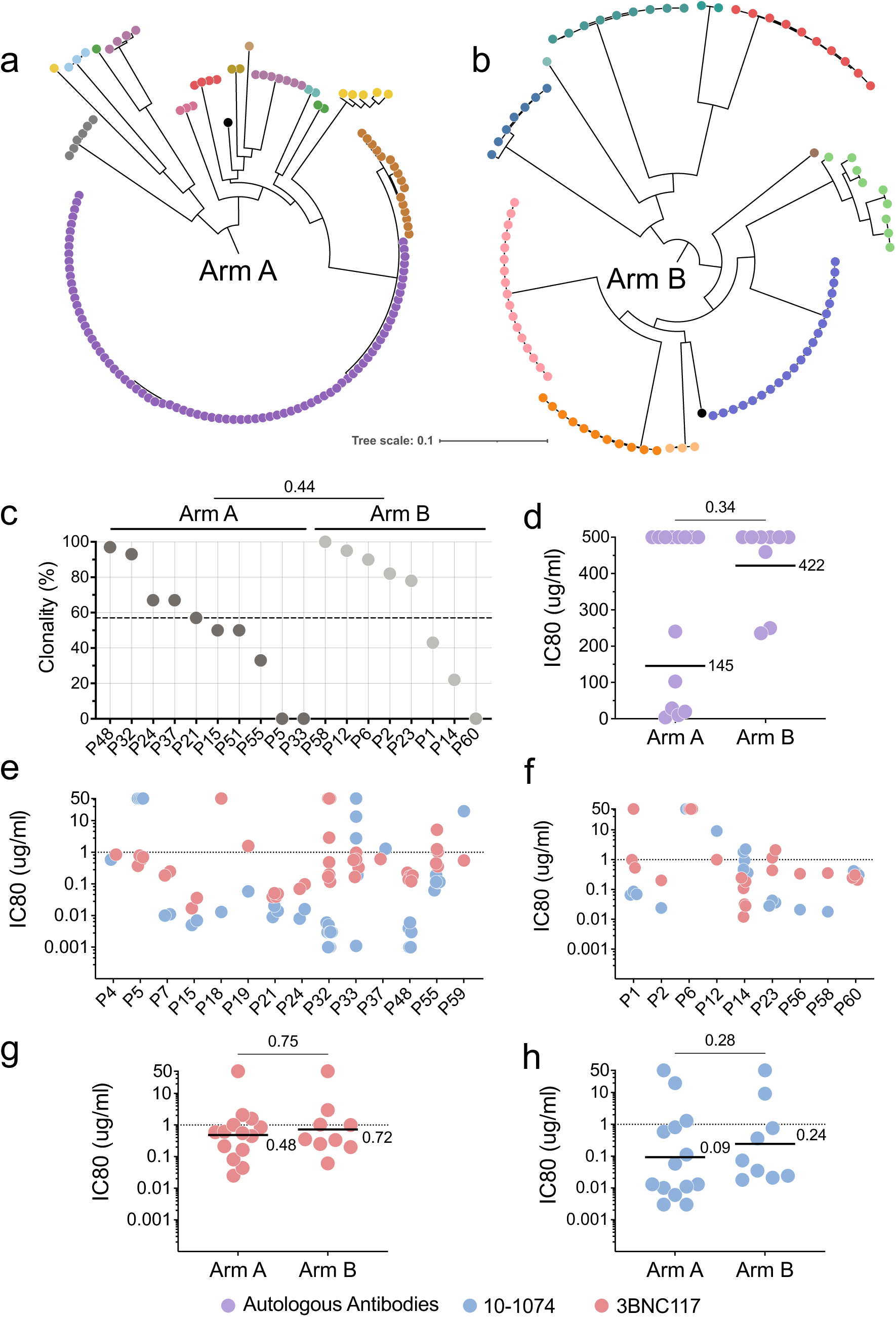
Baseline reservoir characteristics. **a,b**, Circular phylogenetic trees using the generalized time-reversible (GTR) model depict clonal distribution of intact proviral nucleotide sequences based on *env* (**a**) Arm A and (**b**) Arm B. Colors represent different participants. **c**, Intact reservoir clonality for the indicated participants, calculated based on amino acid sequences of *env* for individuals with ≥3 intact proviral sequences. Dashed line indicates combined average clonality of both Arms. **d**, Baseline reservoir sensitivity to pre-infusion autologous IgG antibodies, expressed as IC80; each dot represents the geometric mean sensitivity of proviruses obtained from one individual. Line and numbers show overall geometric mean for all participants in Arms A (n=13) and B (n=9). **e,f**, Baseline reservoir neutralization sensitivities to 3BNC117 (red) and 10-1074 (blue), expressed as IC80. Dotted lines indicate 1 mg ml^-1^. **e**, Individual pseudovirus IC80s for the indicated participants in Arm A. **f**, Individual pseudovirus IC80s for the indicated participants in Arm B. **g,h**, Each dot represents the geometric mean IC80s for all pseudoviruses from an individual participant against (**g**) 3BNC117 and (**h**) 10-1074. Arm A n=14 and Arm B n=9. Group comparisons in **c,d** and **g,h** were performed using the Mann–Whitney test, with statistical significance defined as *P* < 0.05.

To determine whether baseline reservoir size was associated with virologic outcomes after ATI, we compared intact proviral reservoir measurements with time to ART restart and peak viremia. There was no significant correlation between the size of the intact proviral reservoir and time to ART restart in the placebo or bNAb arms of the study, as measured by either ddPCR or Q4PCR (Extended Data Fig. 3). In addition, there was no correlation between the reservoir size and peak viremia prior to ART restart in either group (Extended Data Fig. 3).

Sixty-five pseudotype viruses from distinct intact reservoir-derived *env* sequences from 14 and 9 individuals in Arms A and B, respectively, were tested for neutralization sensitivity to autologous baseline IgG, 3BNC117, and 10-1074 in TZM-bl assays ^47^ (Fig. 1d-h and Supplementary Tables 3 and 5). Consistent with early therapeutic intervention, only 8 of these 23 individuals showed autologous antibody neutralizing activity with an IC80s <250 ug ml^-1^ of purified IgGs (corresponding to ≅1:100 dilution of serum) against their reservoir proviruses (Fig. 1d, and Supplementary Tables 3 and 5). To assess whether autologous neutralizing activity changed over time, we performed longitudinal neutralization assays following depletion of residual infused bNAbs (see methods). Autologous IgG was tested both against baseline reservoir-derived viruses and against a panel of HIV strains resistant to 3BNC117 and 10-1074 to evaluate neutralization breadth. We found no increase in autologous neutralizing activity against baseline reservoir viruses and no measurable expansion of neutralization breadth in samples collected at a mean of 68 weeks after ATI from individuals in Arm A (Extended Data Fig. 4).

Reservoir sensitivity to each of the two bNAbs, ranged from an IC80s of <0.001 to >50 ug ml^-1^ for 10-1074 and 0.012 to >50 ug ml^-1^ for 3BNC117, with no significant differences between the two study arms (Fig. 1 e-h). The geometric mean IC80s were 0.48 ug ml^-1^ and 0.72 ug ml^-1^ for 3BNC117 and 0.09 and 0.24 for 10-1074 for Arms A and B, respectively (Fig. 1g and h and Supplementary Table 5). All individuals in the bNAb arm showed reservoir proviral sensitivity to at least 1 of the 2 bNAbs (Supplementary Tables 3 and 5).

### Rebound viremia

We were able to obtain 323 and 370 rebound viral *env* sequences by single genome amplification, from 19 and 16 individuals who experienced viral rebound in Arms A and B, respectively (Supplementary Table 4). To examine the nature of rebound viremia and its relationship to the proviral reservoir, we compared reservoir and rebound *env* sequences from 18 individuals from whom we were able to obtain both intact reservoir and rebound sequences (Fig. 2a and b). Pseudotype viruses obtained from participants in Arms A (96) and B (119) were produced and tested for sensitivity to the 2 bNAbs and to autologous antibodies obtained at the time of entry (Fig. 2c – f, Fig. 3a-f and Supplementary Table 4).

**Figure 2.**
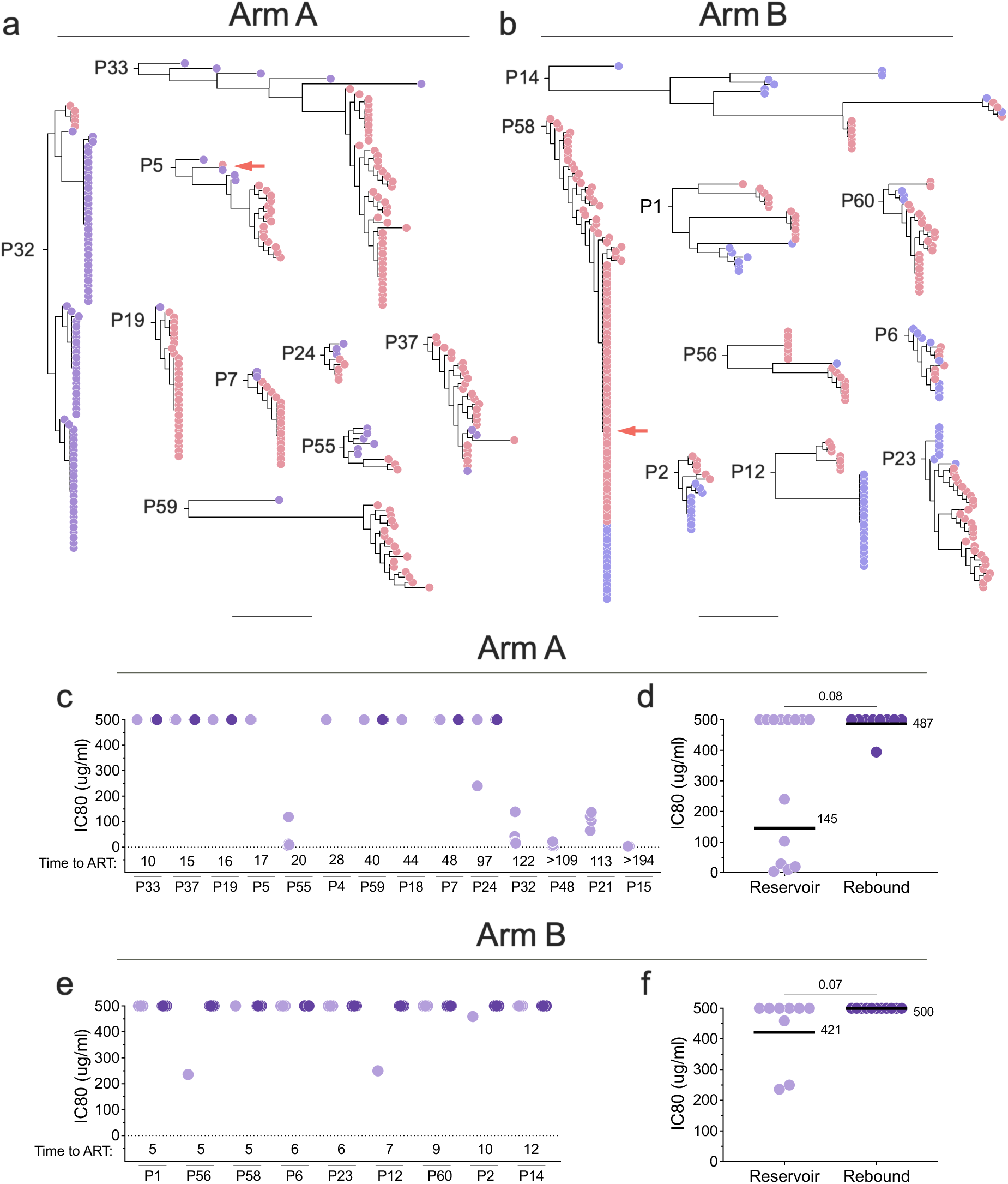
Relationship between intact proviral reservoir and rebound *env* nucleotide sequences from participants in both study arms. **a,b**, Phylogenetic trees were constructed using the generalized time-reversible (GTR) model for participants in Arm A and Arm B. Intact reservoir proviruses are shown in purple and rebound in red. Red arrows indicate sequences shared between reservoir and rebound. **c-f**, Neutralization of reservoir and rebound pseudoviruses by autologous antibodies in participants from Arm A and Arm B. (**c, e**) Individual pseudovirus IC80s. Numbers at the bottom indicate the time to ART restart in weeks. (**d, f**) Each dot represents the geometric mean IC80 for all pseudoviruses from one participant. Arm A reservoir n=13 and rebound n=9. Arm B reservoir n=9 and rebound n=11. Line and numbers indicate geometric mean for all participants. Group comparisons in (**d**) and (**f**) were performed using the Mann–Whitney test, with statistical significance defined as *P* < 0.05.

**Figure 3.**
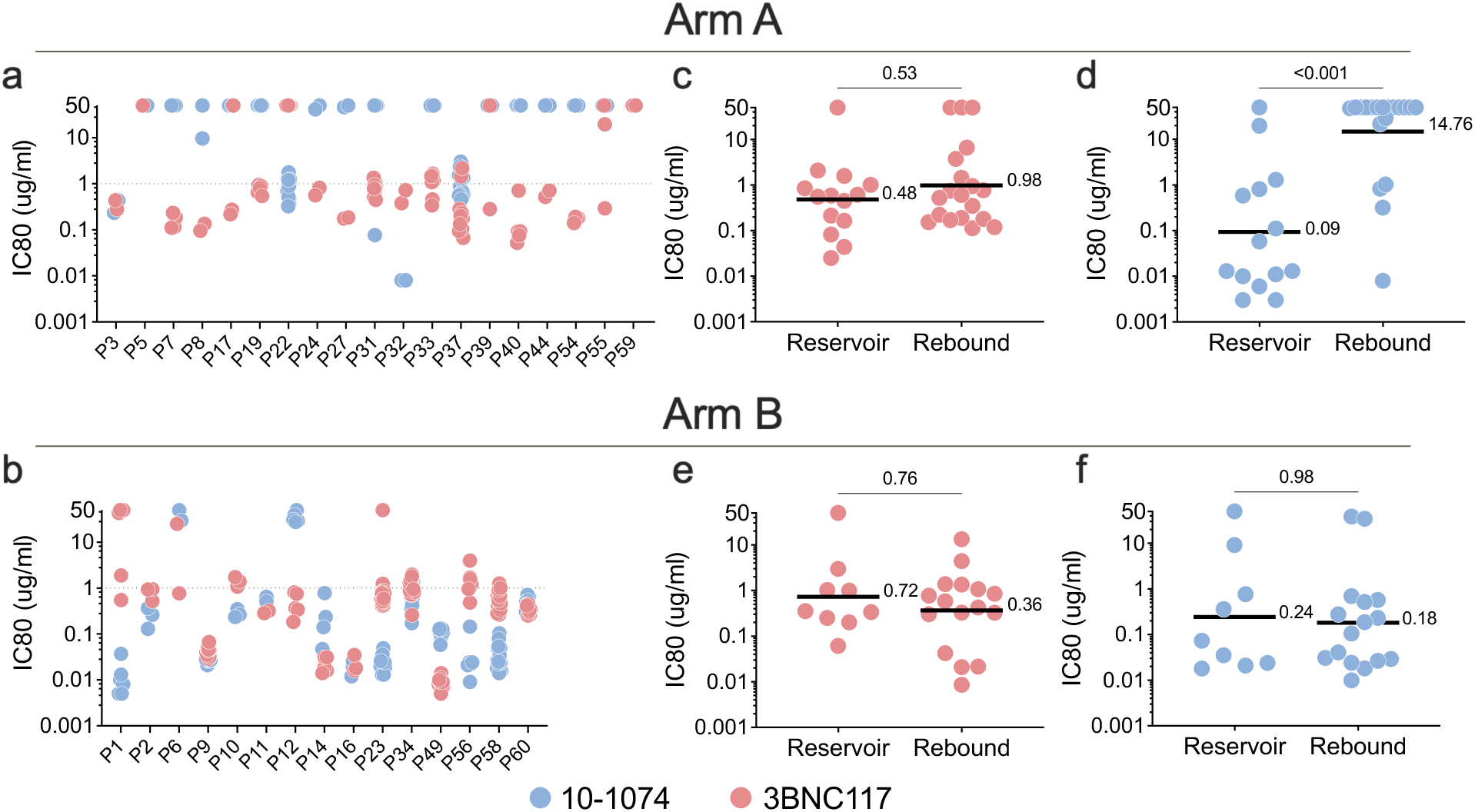
Pseudovirus neutralization by 3BNC117 (red) and 10-1074 (blue) for participants expressed as IC80. **a,b**, Each dot represents an individual rebound pseudovirus for Arm A and Arm B. **c-f**, Each dot represents the geometric mean IC80 for reservoir- and rebound pseudoviruses in one participant Arm A: (**c**) 3BNC117 and (**d**) 10-1074 (reservoir n=14, rebound n=19), and Arm B: (**e**) 3BNC117 and (**f**) 10-1074 (reservoir n=9, rebound n=16). Line and numbers indicate geometric mean for all participants. Group comparisons in (**c,d**) and (**e,f**) were performed using the Mann–Whitney test, with statistical significance defined as *P* < 0.05.

Rebound virus sequences rarely overlap with the dominant latent intact proviral clones found in the circulating reservoir ^48,49^. This phenomenon remains poorly understood but has been attributed in part to suppression of reservoir viruses by autologous antibodies ^35,50^. Among 33 intact latent proviral reservoir clones (91 sequences) obtained from 9 control arm participants, we found only 2 that overlapped with rebound sequences (Fig. 2b). There was no overlap between the reservoir and rebound sequences in the remaining 7 control participants, only 3 of whom had measurable levels of autologous neutralizing antibodies to the reservoir (Fig. 2e and Supplementary Tables 3 and 5). Thus, autologous antibody neutralizing activity cannot entirely account for the disparity between intact proviruses found in the reservoir and the dominant viruses found at the time of rebound in this study of individuals receiving early ART therapy. Nevertheless, rebound viruses in the 3 individuals from the control group that had measurable autologous neutralizing antibodies to the reservoir were resistant to their own antibodies (Fig. 2d-f). As expected, when autologous antibodies are present at baseline (during ART), rebound viruses are enriched in variants that are autologous antibody resistant ^33,34^.

Pseudotype viruses obtained from rebound were also tested for sensitivity to the 2 bNAbs (Fig. 3a-f). Consistent with the absence of selective pressure in the control arm, the geometric mean bNAb sensitivity of all reservoir and rebound pseudotype viruses obtained from Arm B participants was not significantly different for either antibody (3BNC117 p=0.76, 10-1074 p=0.98, Fig. 3b, e and f).

In contrast, selection for bNAb resistance was evident among rebound viruses obtained from Arm A participants (Fig. 3a, c and d). The geometric mean neutralizing IC80s for 10-1074 against reservoir and rebound pseudoviruses was 0.09 and 14.7 mg ml^-1^, respectively with the majority of the rebound viruses being completely resistant (IC80 > 50 ug ml^-1^) (P<0.001 Fig. 3d and Extended Data Fig. 5b). In contrast, there was a much smaller difference in the geometric mean IC80s between reservoir and rebound for 3BNC117 that did not reach statistical significance (p=0.53, Fig. 3c and Extended Data Fig. 5a). Only 3 of 19 participants that received bNAbs showed rebound viruses that were uniformly resistant to 3BNC117 (Fig. 3a and Supplementary Tables 6 and 7). In conclusion, 10-1074 escape variants were far more likely to emerge than 3BNC117 resistant variants during dual bNAb therapy, as reported in earlier studies ^25,27,51^.

### Viral Control and Reservoir Antibody Sensitivity

To determine whether reservoir sensitivity to either autologous antibodies or bNAbs was associated with control of viremia, we compared baseline autologous neutralizing titers and bNAb IC80s to reservoir viruses and time of ART restart (Fig. 4 a-g). These analyses were limited to participants from whom reservoir viral sequences could be obtained. The mean time to ART restart among the three individuals in the control group that exhibited autologous antibody neutralizing activity against reservoir viruses was 7.3 compared to 8.5 weeks to all participants (Fig. 4b). Thus, in this small sample we see no measurable effect of the autologous antibodies in the absence of bNAb therapy. In contrast, the mean time to ART restart in the six bNAb recipients that showed baseline autologous neutralizing activity was 108 weeks compared to 27.5 to the 13 that did not (p=0.008, Fig. 4a). Consistent with this observation, there was a significant correlation between pre-infusion autologous neutralizing antibody titers to reservoir viruses and time to ART restart for individuals in the bNAb arm (n=14, r=-0.69 p=0.004, Fig. 4c). Time to ART restart was also correlated with initial reservoir proviral sensitivity to 10-1074 (r=-0.79, P<0.001) but there was only a trend for 3BNC117 (r=-0.45, p=0.06, Fig. 4d and e). Although we were unable to obtain additional intact pro-viral reservoir sequences, we were able to obtain intact *envs* which were used to construct pseudotype viruses that were tested for sensitivity (n = 5 Arm A) (Supplementary Table 3). When combined with the data obtained from intact proviruses, the correlation with time to ART restart remained significant for 10-1074 and autologous antibodies and became significant for 3BNC117 (Extended Data Fig. 6). There were no consistent associations between baseline autologous antibodies and baseline HIV-specific CD4+ or CD8+ T-cell responses ^52^ (Extended Data Fig. 7). While correlations were observed for individual assays (AIM CD4 response to Nef and CD8 CTV response to Nef), these were not consistent across assays and sample size was limited precluding robust comparisons (Extended Data Fig. 7).

**Figure 4.**
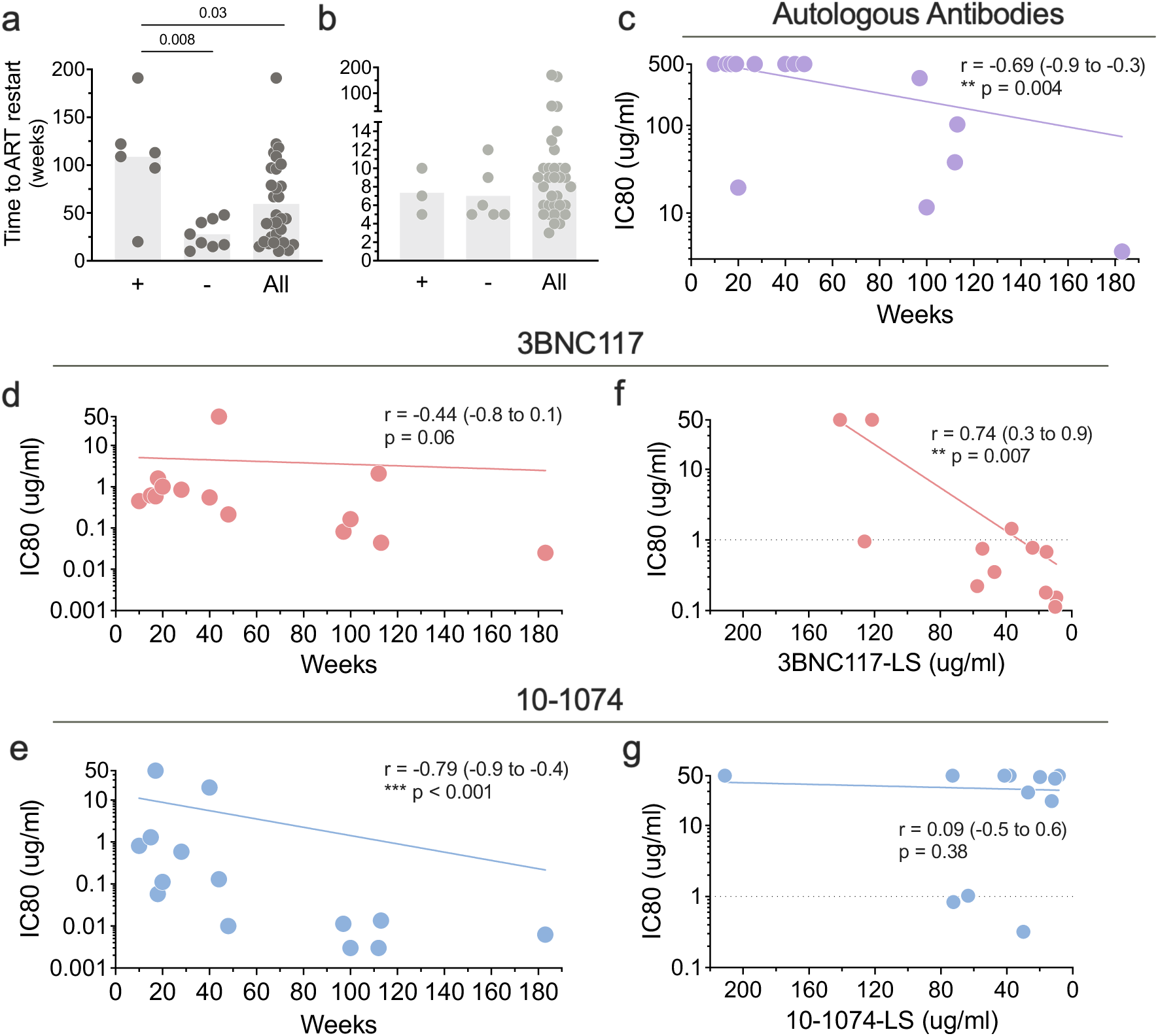
Viral suppression and reservoir sensitivity to autologous antibodies, 3BNC117 and 10-1074. **a,b**, Time to ART restart for individuals that did (+) or did not (-) display reservoir sensitivity to autologous antibodies or all participants. (**a**) Arm A and (**b**) Arm B. Each dot represents the time to rebound for an individual participant. The bars represent the geometric mean for each group. Mann–Whitney group comparisons, with \**P* < 0.05, *\*\*P* < 0.01, *\*\*\*P* < 0.001 considered significant. **c-e**, Correlation between time to ART restart and baseline reservoir sensitivity to (**c**) autologous antibodies, (**d**) 3BNC117, and (**e**) 10-1074. Each dot represents the geometric mean IC80 (Y axis) and time to rebound in weeks (X axis) for one participant. **f,g**, Correlation between IC80 from rebounding viruses for (**f**) 3BNC117 or (**g**) 10-1074 and the bNAb concentration at the time of rebound. Each dot represents the geometric mean IC80 (Y axis) and bNAb concentration at the time of rebound in micrograms per milliliter (X axis) for one participant. Nonlinear curve fitting was performed with *x* as a linear function of time or bNAb concentration, and *y* as a logarithmic function of IC80 values. Correlation coefficients were calculated using Spearman’s *r* with 95% confidence intervals.

Although potentially insufficient on their own, persistent low levels of the 2 long-acting antibodies could contribute to control. The mean concentration of 3BNC117-LS and 10-1074-LS at 96-123 weeks after bNAb infusion in 7 Arm A controllers was 0.70 and 0.96 ug ml^-1^, respectively (Supplementary Table 6). To evaluate the combined influence of endogenous and therapeutic antibody pressure on rebound in Arm A participants, we performed time-to-event analyses, modeling time to ART restart as a function of baseline reservoir sensitivity to autologous antibodies, 10-1074, and 3BNC117 using IC80 values. In univariable analyses, resistance to both autologous antibodies and 10-1074 was associated with shorter time to ART restart. Because reservoir sensitivity to autologous antibodies tended to positively correlate with sensitivity to 10-1074 (and to a lesser extend 3BNC117), these variables were partially collinear and may reflect overlapping antiviral susceptibility profiles. Consistent with this, the associations were attenuated in exploratory multivariable models when included together (Supplementary Table 7). Notably, autologous antibodies and 10-1074 retained significance when adjusted for 3BNC117 alone.

There was an inverse correlation between 3BNC117 concentration at the time of rebound and the IC80 of the rebounding virus to the bNAb (Fig. 4f). In contrast, this relationship was not observed for 10-1074, which displayed a broad range of concentrations at rebound (Fig. 4g). These results are consistent with the idea that individuals that rebound do so when they develop resistance to 10-1074 and have subtherapeutic concentrations of 3BNC117 in circulation. Although the number of reservoir viruses analyzed was a small fraction of the total present in any given individual, the data indicates that the probability of escape from 10-1074 was directly related to the sensitivity of the intact proviruses in the initial reservoir (Fig. 4e). Furthermore, baseline reservoir size did not correlate with the presence or magnitude of autologous neutralizing activity against reservoir viruses, indicating that delayed rebound in bNAb-treated participants was not explained simply by a smaller baseline reservoir (Extended Data Fig. 8)

Among the 29 Arm A participants that remained in follow up according to protocol, 13 went back on ART before meeting virological restart criteria (2 consecutive VLs ≥10^5^ HIV-1 copies ml^-1^ or 6 consecutive VLs ≥10^3^, Extended Data Fig. 9). In a post hoc subgroup analysis, of the 16 remaining, 11 (68%) maintained fluctuating viremia and did not meet ART re-start criteria for 16 to >58 weeks (Fig. 5a). In contrast, only 2 of the Arm B control showed prolonged fluctuating viremia ^37^. We were able to obtain viruses from 7 participants from time points associated with fluctuating viremia, and all were resistant to pre-infusion autologous antibodies when present. In addition, 5 of these individuals showed viruses that were highly resistant to 10-1074, but retained sensitivity to 3BNC117 (Fig. 5c and d). These findings identify fluctuating viremia as a distinct post-bNAb virologic phenotype that others have recently associated with enhanced T cell immunity ^24,29,31^.

**Figure 5.**
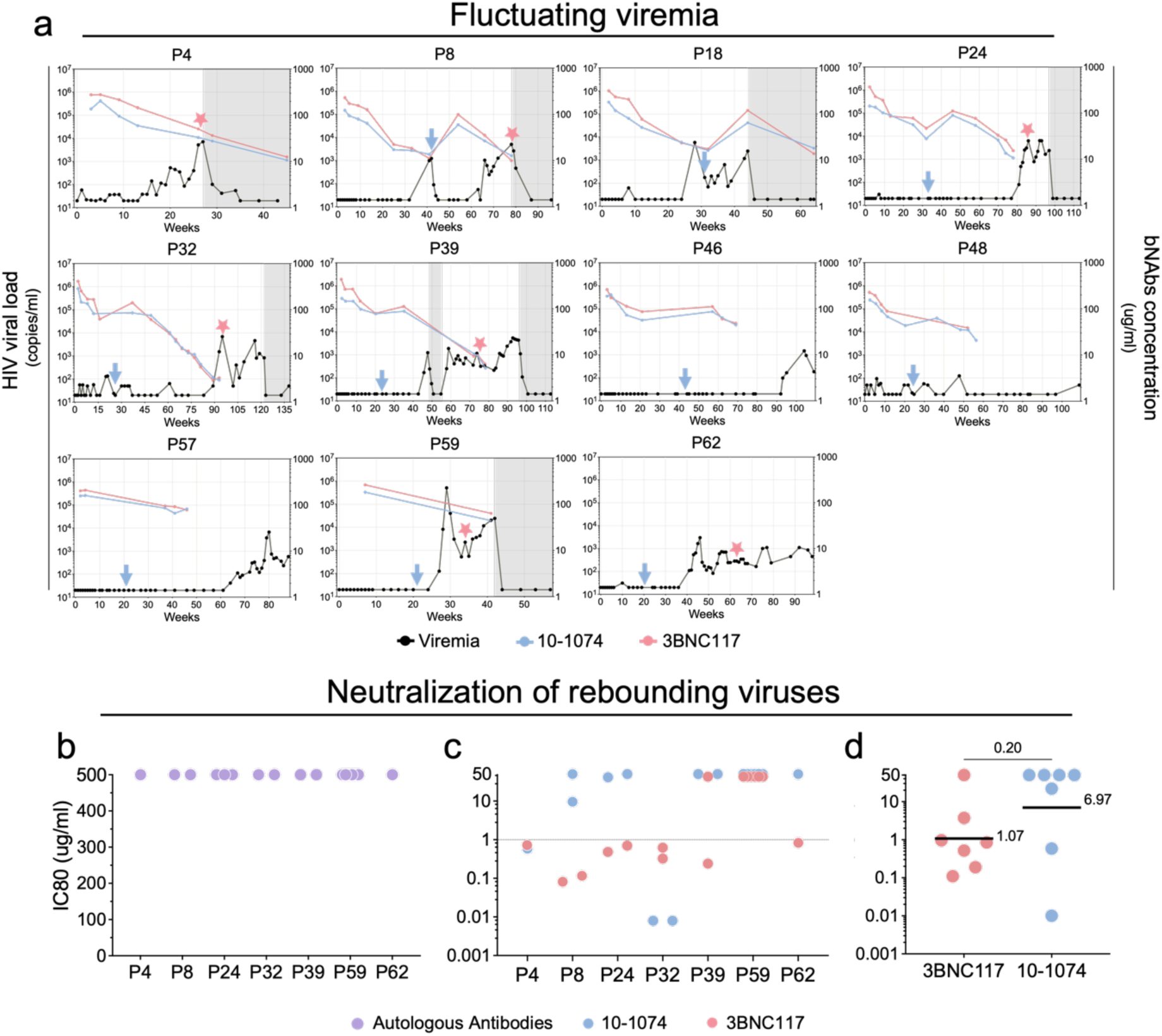
Virologic dynamics and antibody sensitivity during fluctuating viremia following bNAb therapy. **a,** Longitudinal plasma HIV-1 RNA levels (black) and circulating bNAb concentrations for 10-1074 (blue) and 3BNC117 (red) are shown for individual participants who maintained fluctuating viremia (defined as >16 weeks without sustained HIV-1 RNA >1,000 copies ml^-1^). Time is shown in weeks on the x axis, with HIV-1 RNA copies per milliliter and bNAb concentrations in micrograms per milliliter plotted on log10 scales on the y axes. Blue arrows indicate administration of a second bNAb infusion. Gray shading denotes periods on antiretroviral therapy. **b,** Neutralization sensitivity (IC80) of rebound-derived pseudoviruses to pre-infusion autologous IgG for individual pseudoviruses. **c,** Neutralization sensitivity (IC80) of rebound-derived pseudoviruses to 3BNC117 (red) and 10-1074 (blue), shown for individual pseudoviruses. **d,** Geometric mean IC80 values per participant comparing sensitivity to 3BNC117 and 10-1074. Lines and numbers indicate geometric means across participants. Group comparisons were performed using the Mann–Whitney test, with statistical significance defined as *P* < 0.05.

### Reservoir dynamics

To determine how the bNAbs impact the size of the proviral reservoir in Arm A, we compared the intact and defective proviral reservoirs at baseline with pre-rebound samples collected at a mean of 52 weeks after the initial infusion (n=22, Fig. 6 and Supplementary Table 2). Reservoir size was also measured in Arm B after a mean of 52 weeks directly before the second treatment interruption ^37^. We used ddPCR for these measurements because the relatively small reservoir size limited the number of samples from which we were able to obtain paired Q4PCR measurements (Supplementary Table 2). Although, there was a great deal of variation between individuals in Arm A over time, the intact reservoir but not the defective was significantly smaller after bNAb therapy with an estimated half-life of 36 weeks (n=22, Fig. 6a and Supplementary data Fig. 10). In contrast, there was no measurable change in reservoir size in Arm B (n=20, Fig. 6b). As expected from the relatively small overall change in the intact reservoir in Arm A, compared to estimated reservoirs of millions of cells, there was no correlation between the change in reservoir size and time to ART restart (Extended Data Fig. 11).

**Figure 6.**
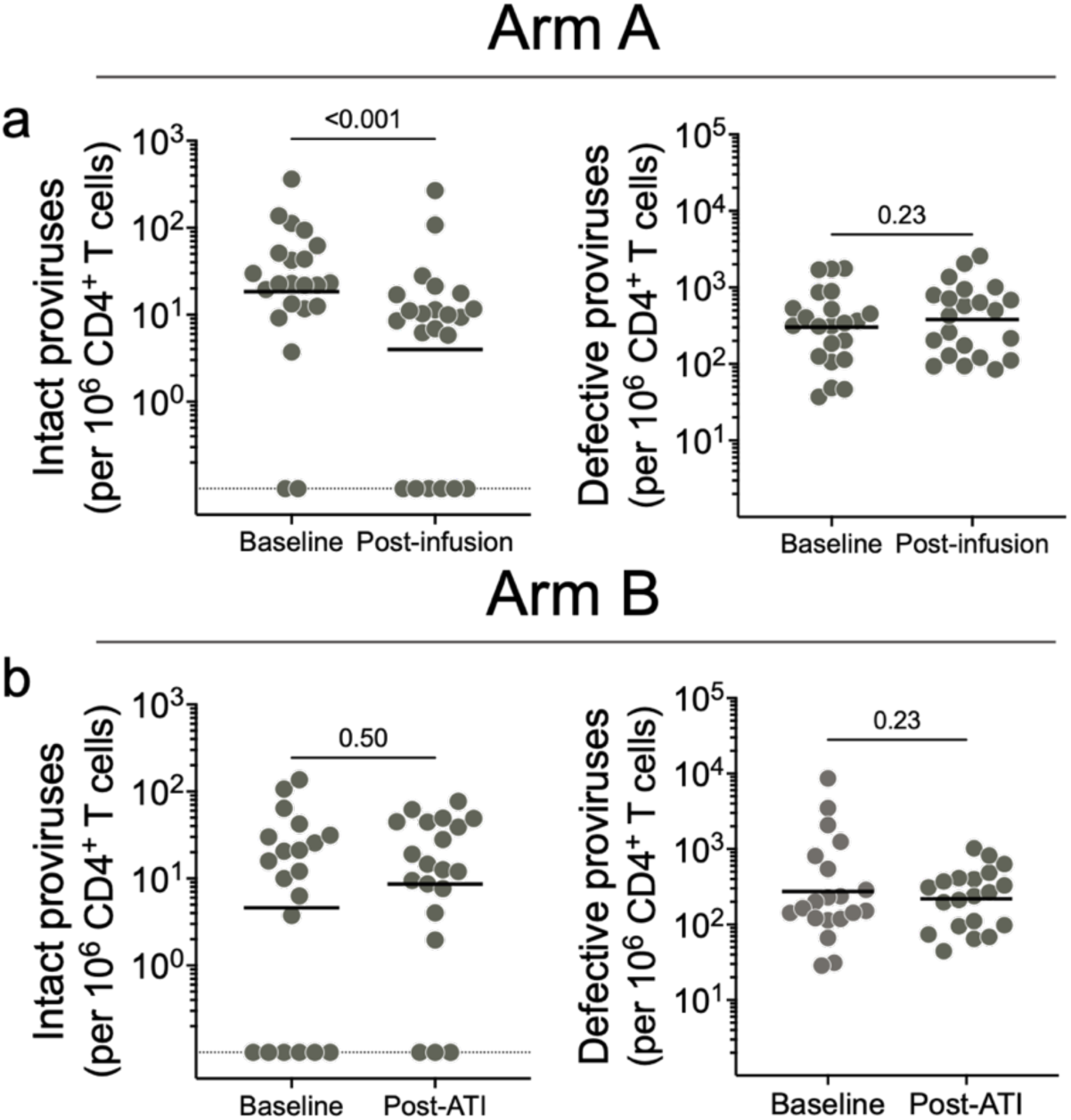
Intact and defective proviral reservoirs measured by ddPCR. **a,b,** Quantification of intact and defective proviral reservoirs by digital droplet PCR at baseline and at follow-up timepoints in participants from Arm A (n = 22) and Arm B (n = 20). Changes over time within each group were evaluated using the Wilcoxon matched-pairs signed-rank test (P < 0.05).

## Discussion

RIO is an ongoing double-blind, randomized placebo-controlled human trial in which participants that started ART during primary or early infection were randomly assigned to a group that received long acting LS variants of 3BNC117 and 10-1074, Arm A, and a control group, Arm B. Both groups subsequently discontinued ART ^37^. Upon rebound participants in Arm B received ART and the 2 bNAbs before undergoing a second treatment interruption ^37^. Despite similar pre-infusion reservoir measurements, the groups differed significantly in time to rebound ^37^, and in the nature of the rebounding viruses. After 96 weeks 25% and 6% of the individuals in the active and control arms, respectively, remained off ART. Viral rebound occurred across a wide temporal range in Arm A encompassing periods of high circulating bNAb concentrations as well as later timepoints when antibody levels were low. Because participants received the LS versions of the bNAbs, antibody exposure extended over long periods of time during ATI. Prolonged control was associated with pre-existing autologous neutralizing antibodies and sensitivity to 10-1074. Paired pre-infusion and pre-rebound intact reservoir measurements showed a decrease in intact, but not defective, proviruses in Arm A that was not observed in Arm B participants. Notably, participants in Arm B experienced viral rebound and periods of detectable viremia between sampling time points, which may have contributed to increase of the circulating reservoir. In addition, differences in baseline intact reservoir between the two arms may further confound these comparisons. Therefore, these associations are correlative and based on a limited number of individuals, but the observed reservoir decay is suggestive of altered reservoir dynamics in this cohort of PLWH undergoing ATI under LS-bNAb exposure.

As expected, based on their relatively early start on therapy, participants in RIO had small HIV-1 proviral reservoirs ^41–43^. Nevertheless, most of the intact reservoir in participants was clonal and did not contribute to rebound ^48,49^. Why reservoir proviruses are rarely found among rebound viruses is not entirely understood, but there is an inverse correlation between the size of an expanded clone and the probability that the sequence will be found among rebound viruses ^12^. The difference has been attributed to selection for proviral integration into silent sites in the genome ^18–20,53,54^, to autologous antibody mediated suppression ^35,50^, and the possibility that rebound arises from tissue reservoirs not sampled in peripheral blood, as suggested by non-human primate studies ^55,56^. Notably, the disparity between reservoir and rebound viruses was evident in control arm individuals that did not have measurable autologous antibodies or receive bNAbs. Thus, the discrepancy between reservoir proviruses in circulation and rebound viruses cannot entirely be explained by autologous antibody suppression of viremia in this cohort of early treated individuals.

HIV-1 infection is associated with the development of antibodies that neutralize the autologous virus and continually select for viruses that express resistant variants during the course of the infection ^33,34^. Moreover, autologous antibodies can sieve the outgrowth of reservoir viruses *in vitro* ^35,50^. But although autologous antibodies alter the viral swarm by selection of resistant variants, they fail to control viremia, leading to the suggestion that antibodies are ineffective against HIV-1. This idea was further supported by the observation that passively administered first generation bNAbs had little, if any, measurable effect on infection ^57–59^. However, subsequent experiments in mice and macaques showed that administration of broader and more potent bNAbs suppresses viremia and elicits CD8+ T cell responses that lead to prolonged control of infection ^21–23,60–62^. In addition, small interventional studies in which people living with HIV-1 received second generation bNAbs demonstrated that antibodies can be both safe and effective in reducing viremia in the absence of ART ^28,31,51,63–65^. Moreover, a fraction of individuals receiving bNAbs during treatment interruption appear to control the infection for an unexpectedly long time.

Enhanced CD8+ T cell responses were associated with bNAb therapy in some but not all reported clinical studies and therefore other mechanisms are thought to contribute to enhanced control ^24,30,31^. The RIO trial is a blinded placebo-controlled trial in which the data indicate that individuals treated early who have pre-existing autologous neutralizing antibodies are more likely to control for longer periods of time. Although, the association between autologous antibodies and suppression is correlative and does not establish a direct mechanistic role for autologous antibodies in viral suppression, autologous antibodies could function as an adjunct to the 2 bNAbs making it more difficult for the virus to escape from combined antibody pressure ^66^. Notably, sensitivity to autologous antibodies correlated with sensitivity to 10-1074, suggesting partially overlapping aspects of viral neutralization susceptibility ^67,68^. Several mechanisms by which antibodies can contribute to viral control have previously been documented, including direct neutralization of circulating viruses ^69^, clearance of infected cells *in vivo* ^61,70^, and antibody-mediated enhancement of antigen presentation to elicit cellular immune responses ^71–73^. These mechanisms represent a plausible framework by which autologous antibodies could contribute to the delayed rebound observed in a subset of participants.

A significant fraction of the participants in the bNAb arm of the RIO trial maintained fluctuating viremia for 16 to >58 weeks in the face of complete or partial bNAb resistance (11 of 29 or 38% of Arm A). Although it did not reach significance, enhanced post-bNAb control has also been recently reported in the TITAN trial and in a single arm study of 10 individuals that received bNAbs, a TLR agonist and a vaccine ^24,29^. Similar results were also observed in bNAb treated macaques ^21–23^. Together, these observations point to fluctuating viremia as a reproducible post-bNAb virologic phenotype. The precise mechanistic explanation for these observations is not well defined, but the role of residual antibody as one contributing component cannot be dismissed.

Antibody monotherapy therapy like small molecule therapy results in selection for resistant variants. Combination therapy durably suppresses viremia because the probability of developing escape mutations at a combination of non-overlapping different sites is less likely. Consistent with this idea, rebound typically occurred in the RIO trial under functional monotherapy. Rebound viruses were most frequently resistant to 10-1074 and pre-existing autologous antibodies. In contrast, selection against 3BNC117 was far more limited with no significant difference between initial reservoir and rebound sensitivity to the antibody, suggesting that the antibody was sub-therapeutic at rebound. The median estimated 3BNC117-LS concentration at rebound was 52.4 μg ml^-1^ which is similar to the median 3BNC117 IC80 for these participants at the time of rebound (41.8 ug ml^-1^, Fig. 4f), and higher than the estimates in previous small clinical trials ^25,27^. These findings further support the interpretation that fluctuating viremia and delayed rebound may not be explained by antibody effects alone.

RIO trial participants who received bNAbs demonstrated accelerated intact reservoir decay, 0.75 years compared to prior estimates of 4-7 years ^14–17^. However, the accelerated intact reservoir decay was not uniform across the cohort with 85% of the cohort showing a decrease. In contrast, there was no measurable change in reservoir in Arm B between the first and second treatment interruption. The difference could be due to reservoir changes in response to viremia in Arm B, or the addition of ART to bNAb therapy in Arm B or both. Accelerated reservoir decay was also a feature of a small study in which participants received multiple doses of a short acting native version of 3BNC117 and 10-1074 ^25^, but not in other small studies ^24,29^. The difference may be due to inherent variability and the small numbers of participants assayed, who were treated during chronic infection as opposed to early therapy participants in the RIO trial, differences in the antibodies used and their half-lives, and finally significant variability in the nature of the reservoir between individuals. Accordingly, we interpret intact reservoir decay in RIO as suggestive of altered reservoir dynamics under the specific conditions of early treatment and long-acting bNAb exposure. Why some participants in this trial showed only small or no change in reservoir size remains to be determined but may be due to heterogenous and/or variable levels of proviral transcription among CD4+ T cells and even among members of a clone of CD4+ T cells ^74–76^. Moreover, the observed changes in reservoir size cannot account for and do not correlate with prolonged antibody-mediated suppression (Extended Data Fig. 11). Far greater changes in reservoir would be required to alter the time to rebound in most individuals.

In conclusion, the data establishes that a significant fraction of people living with HIV-1 who receive bNAb therapy can partially control viremia for prolonged periods of time. Correlates of prolonged control of HIV-1 after bNAb therapy include pre-existing autologous antibodies and pre-existing or enhanced CD8+ T cell responses ^29,36,52^, suggesting that vaccination to elicit or enhance pre-existing host immunity may further increase the number of individuals exhibiting post-bNAb control.

### Limitations of this Study

Although 68 individuals were recruited into the RIO study, we were unable to obtain sequence information on the HIV-1 reservoir in 33 of the participants, thereby limiting the number of participant samples available for analysis. This constraint primarily reflects technical limitations of single-genome amplification in individuals treated early, who often harbor small circulating reservoirs, as well as issues related to sample availability and non-B clade viruses. Analyses of autologous neutralizing activity were restricted to participants with detectable reservoir-derived viral sequences. Moreover, sampling was limited to peripheral blood and may not entirely reflect tissue reservoirs. This problem was further accentuated when considering the relationship between autologous antibodies and virologic outcomes because early ART interferes with the development of autologous neutralizing antibody responses. Associations between autologous antibodies and delayed rebound are correlative, and reservoir decay estimates are limited by participants with values below assay detection. Finally, the timing of sample acquisition was pre-determined and did not always correspond to timepoints of interest limiting analysis of rebound viremia.

## Methods

### Ethics statement

The RIO trial was approved by the London Research Ethics Committee (REC reference 19/LO/1669), the UK Health Research Authority, and relevant institutional review boards at participating sites. The study was conducted in accordance with the principles of the Declaration of Helsinki and Good Clinical Practice guidelines. All participants provided written informed consent prior to enrollment. The trial was registered at ClinicalTrials.gov (NCT04319367). Trial oversight included independent Data Monitoring and Trial Steering Committees that reviewed trial safety and overall conduct throughout the study.

### Study participants

The RIO trial is a randomized, placebo-controlled, double-blinded phase II study ^38^. Participants were recruited across multiple HIV clinical research sites in the United Kingdom and on site in Denmark. This study enrolled individuals who initiated ART during primary or early HIV infection (defined as nadir CD4 count > 500 cells ml^-1^) and remained on suppressive ART for at least 1 year. Primary infection was defined according to standard clinical and laboratory criteria at the time of diagnosis, as detailed in the RIO clinical protocol. Early-stage infection was operationally defined as ART initiation prior to substantial immune compromise, using a nadir CD4⁺ T-cell count >500 cells/µL as a pragmatic surrogate for early disease stage. Additional key inclusion criteria included age 18–60 years, ability to provide informed consent, willingness to comply with study visits and blood sampling, no evidence of viral insensitivity to either 10-1074-LS or 3BNC117-LS, current CD4⁺ T-cell count >500 cells ml⁻¹ or CD4:CD8 ratio >1.0, adequate haemoglobin, body weight ≥50 kg, and hepatitis B and C screening consistent with protocol eligibility. Major exclusion criteria included significant cardiovascular disease, malignancy except squamous cell skin cancer, opportunistic infection or significant comorbidity, HTLV-1 co-infection, current or planned systemic immunosuppressive therapy, participation in another interventional trial, history of severe reaction to antibody infusion, treatment with IV immunoglobulin or other monoclonal antibodies during the trial, clinically significant abnormal laboratory results, active alcohol or substance use likely to impair adherence, insufficient venous access, pregnancy or breastfeeding, or concern regarding risk-reduction precautions during ATI.

A total of 68 participants were enrolled between May 2021 and July 2024 and were randomly assigned in a 1:1 ratio into two arms as part of a two-stage clinical design. The RIO trial was powered for the primary clinical endpoint as described in the accompanying clinical manuscript^37^. Briefly, the study was designed to detect a reduction in the rebound rate at 36 weeks from 90% in Arm B to 55% in Arm A (hazard ratio 0.35), with 90% power at a 5% significance level. No formal power calculations were performed for the exploratory mechanistic analyses reported here. Sex and gender information were collected by self-report at study enrollment. Race and ethnicity information were self-reported according to local clinical site procedure. Prespecified sex-based analysis were not performed because the study was not powered for these subgroup analyses. Blood samples were collected at baseline and at multiple time points following 3BNC117-LS and 10–1074-LS or placebo infusions. Samples were processed briefly after collection, with serum and plasma stored at −80°C. Peripheral blood mononuclear cells (PBMCs) were isolated by density gradient centrifugation. The PBMC number was determined either manually or using an automated cell counter (Vi-Cell XR; Beckman Coulter), and cells were cryopreserved in liquid nitrogen in fetal bovine serum supplemented with 10% DMSO.

### Definitions of virological control and ART-free status

Virological control was defined as sustained plasma HIV-1 RNA <1,000 copies ml^-1^ during treatment interruption. ART restart criteria were defined per protocol as either (i) two consecutive plasma HIV-1 RNA measurements >10⁵ copies ml^-1^, or (ii) six consecutive plasma HIV-1 RNA measurements >10³ copies ml^-1^. In addition to these virological criteria, ART could be restarted for other predefined clinical or participant-driven endpoints not meeting the above thresholds, including participant preference or clinician judgment. All ART restart events and their classification were adjudicated by an independent Endpoint Adjudication Committee (EAC). The distribution of ART restart outcomes by study arm is summarized in Extended Data Table 1.

### Intact proviral DNA analysis (IPDA)

The frequency of CD4+ T cells harboring intact, 5’-deleted, and 3’-deleted proviruses was determined using a ddPCR assay modified from the Intact Proviral DNA Assay (IPDA) ^10^. Briefly, CD4+ T cells were enriched from PBMCs using a negative immunomagnetic selection kit (Miltenyi Biotec). Genomic DNA was extracted using the QIAamp DNA Mini Kit (Qiagen) according to the manufacturer’s instructions. DNA concentrations were quantified using the Qubit 3.0 Fluorometer with the Qubit dsDNA BR Assay Kit (Thermo Fisher). Samples were from the pre-infusion time point and obtained from an aviremic time from a minimum of six weeks before rebound, when HIV-1 RNA was < 20 copies ml^-1^.

For ddPCR, two sets of primers and probes specific to the HIV Gag and Env regions, along with two fragments of the housekeeping gene RPP30, were utilized in separate reactions ^77^. HIV quantification was performed using 750 ng of genomic DNA per sample with the following primers and probes for the HIV Gag region: Forward (5’-GACTAGCGGAGGCTAGAAGGAGAGA-3’), Reverse (5’-CTAATTCTCCCCCGCTTAATAYTGACG-3’), and Probe (5’-6FAM-ATGGGTGCGAGA-IABkFQ-3’). For the HIV Env region: Forward (5’-AGTGGTGCAGAGAGAAAAAAGAGC-3’), Reverse (5’-GTCTGGCCTGTACCGTCAGC-3’), and Probes (5’-VIC-CCTTGGGTTCTTGGGA-MGB-3’ and an unlabeled hypermutated probe 5’-CCTTAGGTTCTTAGGAGC-MGB-3’). An alternative primer/probe set targeting the PS region was used as a backup: Forward (5’-CAGGACTCGGCTTGCTGAAG-3’), Reverse (5’-GCACCCATCTCTCTCCTTCTAGC-3’), and Probe (5’-6FAM-TTTTGGCGTACTCACCAGT-IABkFQ-3’) ^10^. To measure input cell numbers and correct for DNA shearing, 7.5 ng of DNA was used with RPP30 primers and probes (RPP30-1: Forward: 5′-GATTTGGACCTGCGAGCG-3′, Reverse: 5′-GCGGCTGTCTCCACAAGT-3′, Probe: 5′-6FAM-TTCTGACCTGAAGGCTCTGCGC-IABkFQ-3′; RPP30-2: Forward: 5′-GTGTGAGTCAATCACTAGACAGAA-3′, Reverse: 5′-AAACTGCAACAACATCATAGAGC-3′, Probe: 5′-HEX-AGAGAGCAACTTCTTCAAGGGCCC-IABkFQ-3′). Four technical replicates were performed per sample. Positive and negative controls were included in each reaction.

The ddPCR assays were conducted using the Bio-Rad QX200 AutoDG system with the ddPCR Supermix for Probes (no dUTPs) (Bio-Rad). Thermal cycling conditions included an initial denaturation at 95°C for 10 minutes, followed by 45 cycles of 94°C for 30 seconds and 59°C for 1 minute, with a 2°C/second ramp rate. A final extension was performed at 98°C for 10 minutes, followed by a hold at 12°C. The results were adjusted for DNA shearing using the ratio of double-positive RPP30 partitions and normalized to 10⁶ CD4+ T cells.

### Quadruplex 4-probes real-time quantitative PCR (Q4PCR)

The Q4PCR assay was conducted to characterize the composition of the reservoir, as previously described ^11^. Briefly, total CD4+ T cells were enriched from cryopreserved PBMCs using a negative immunomagnetic selection kit (Miltenyi Biotec). Genomic DNA was extracted from CD4+ T cells using the Gentra Puregene Cell Kit (Qiagen), and DNA concentration was measured using the Qubit dsDNA BR Assay Kit (Thermo Fisher Scientific).

A total range of 1 to 10 x 10^6^ CD4 T cells were screened per participant. An initial outer PCR (NFL1) was performed on genomic DNA at a single-copy dilution using the outer primers BLOuterF (5′-AAATCTCTAGCAGTGGCGCCCGAACAG-3′) and BLOuterR (5′-TGAGGGATCTCTAGTTACCAGAGTC-3′) ^78^. A 1-µl aliquot of the undiluted NFL1 PCR product was then subjected to a Q4PCR reaction, utilizing four primer–probe sets targeting conserved regions of the HIV-1 genome. PS forward, 5′-TCTCTCGACGCAGGACTC-3′; reverse, 5′-TCTAGCCTCCGCTAGTCAAA-3′; probe, 5′-/Cy5/TTTGGCGTA/TAO/CTCACCAGTCGCC-3′/IAbRQSp; env forward, 5′-AGTGGTGCAGAGAGAAAAAAGAGC-3′; reverse, 5′-GTCTGGCCTGTACCGTCAGC-3′; probe, 5′-/VIC/CCTTGGGTTCTTGGGA-3′/MGB; gag forward, 5′-ATGTTTTCAGCATTATCAGAAGGA-3′; reverse, 5′-TGCTTGATGTCCCCCCACT-3′; probe, 5′-/6-FAM/CCACCCCAC/ZEN/AAGATTTAAACACCATGCTAA-3′/ IABkFQ; and pol forward, 5′-GCACTTTAAATTTTCCCATTAGTCCTA-3′; reverse, 5′-CAAATTTCTACTAATGCTTTTATTTTTTC-3′; probe, 5′-/NED/AAGCCAGGAATGGATGGCC-3′/ MGB. Quantitative PCR (qPCR) was performed under the following thermal cycling conditions: an initial denaturation at 94 °C for 10 minutes, followed by 40 cycles of 94 °C for 15 seconds and 60 °C for 60 seconds. qPCR assays were conducted in a 384-well plate format using the Applied Biosystem QuantStudio 6 or 7 Flex real-time PCR system. Data analysis was performed using ThermoFisher Design and Analysis Software 2.4.3. Samples that exhibited reactivity with two or more of the four probes were selected for a nested PCR (NFL2).

The NFL2 reaction was performed on undiluted 1-µl aliquots of the NFL1 PCR product. Each reaction was conducted in a 20-µl volume using Platinum Taq high-fidelity polymerase (Thermo Fisher Scientific) and the primers 3LTRi (5′-TCAAGGCAAGCTTTATTGAGGCTTAA-3′) and U5-638F (5′-GCGCCCGAACAGGGACYTGAAARCGAAAG-3′) ^19^. The thermocycler conditions for NFL2 were the same as those used for the NFL1 PCR. Library preparation and sequencing were conducted as previously described ^11^.

### HIV-1 sequence assembly and annotation

HIV-1 genome reconstruction was performed using an in-house pipeline, Defective and Intact HIV Genome Assembler (DIHIVA), designed for assembling raw sequencing reads into annotated HIV genomes ^25^. The pipeline includes rigorous quality control steps to enhance accuracy and reliability. First, quality control checks were conducted to remove PCR-amplified reads and correct sequencing errors using clumpify.sh from the BBtools package v38.72 (http://sourceforge.net/projects/bbmap). Next, adapter sequences and low-quality bases were trimmed, and potential contaminant reads were removed using the Trim Galore package v0.6.4 (https://github.com/FelixKrueger/TrimGalore). HIV-1 sequences were assembled using SPAdes v3.13.0, and the longest assembled contig was aligned to the HXB2 HIV-1 reference genome via BLAST ^79^. Sequences exhibiting double peaks—regions indicating the presence of two or more viral variants within a sample (defined by a consensus identity cut-off of <70% for any residue)—or samples with an insufficient number of sequencing reads (≤500 reads) were excluded from downstream analyses.

Sequences that did not meet the double-peak criteria (consensus identity for any residue <70%) were further classified as either intact or defective proviruses. Only intact HIV-1 env sequences were considered for subsequent analyses.

### Single-genome amplification of plasma rebound virus env genes

Sequencing of HIV-1 plasma rebound *env* genes was performed as previously described ^80^. In brief, HIV-1 RNA was extracted from viremic plasma samples using the MinElute Virus Spin kit (Qiagen) according to manufactures recommendation. First-strand cDNA synthesis was carried out using SuperScript III reverse transcriptase (Invitrogen) and an antisense primer, envB3out (5′-TTGCTACTTGTGATTGCTCCATGT-3′) for subtype B or OFM19 (5′–GCACTCAAGGCAAGCTTTATTGAGGCTTA-3′) for subtype C. To ensure single-genome amplification, cDNA was endpoint diluted according to Poisson distribution, achieving <30% of wells yielding a PCR product. The envelope gene was then PCR-amplified using subtype B primers envB3out and envB5out (5′-TAGGCATCTCCTATGGCAGGAAGAAG-3′) or subtype C primers OFM19 and Vif1 (5′-GGTTTATTACAGGGACAGCAGAG-3′). A second round of PCR was performed using 1 µl of the first-round PCR product as a template, with subtype B primers envB3in (5′-GTCTCGAGATACTGCTCCCACCC-3′) and envB5in (5′-TAGGCATCTCCTATGGCAGGAAGAAG-3′), or subtype C primers ENV A (5′-GGCTTAGGCATCTCCTATGGCAGGAAGAA-3′) and ENV N (5′-CTGCCAATCAGGGAAGTAGCCTTGTGT-3′). PCR amplifications were conducted using High Fidelity Platinum Taq (Invitrogen) under the following conditions: an initial denaturation at 94°C for 2 minutes, followed by 35 cycles of 94°C for 15 seconds, 55°C for 30 seconds, and 68°C for 4 minutes, with a final extension at 68°C for 10 minutes. PCR products of the expected size were subjected to library preparation and sequencing using the Illumina MiSeq platform.

### Sequence and phylogenetic analysis

Nucleotide alignments of intact env sequences were performed using Muscle v5.1 with PPP algorithm ^81^. Sequences containing premature stop codons, truncations, or frameshift mutations were excluded from further analyses. Maximum likelihood phylogenetic trees were generated from these alignments using FastTree 2.1.11 with the GTR model and 1,000 bootstrap replicates ^82^.

### Pseudotyped-virus production

Selected single-genome sequences from CD4+ T cell reservoir or plasma rebound samples were used as templates to produce pseudoviruses from a CMV promoter as described ^83^. The cytomegalovirus (CMV) promoter was amplified by PCR from pcDNA 3.1 (Life Technologies) using the primers CMVenv (5′-AGTAATCAATTACGGGGTCATTAGTTCAT-3′) and CMVenv1A (5′-CATAGGAGATGCCTAAGCCGGTGGAGCTCTGCTTATATAGACCTC-3′).

Thermocycling conditions were: 94°C for 2 minutes; 30 cycles of 94°C for 30 seconds, 55°C for 30 seconds, and 68°C for 4 minutes. A 1-µl aliquot of the second-round PCR product from NFL amplification (reservoir) or SGA amplification (rebound) was used as a template for env to which we added CMV overhanging regions using the forward primer ENVfwd (5′-CACCGGCTTAGGCATCTCCTATGGCAGGAAGAA-3′) and the reverse primer envB3in or ENV N, depending on the subtype. The CMV promoter amplicon was fused to individual env genes via overlapping PCR with 10 ng of env and 0.5 ng of CMV using CMVenv primer and envB3in or ENV N as the reverse primer. Thermocycling conditions were: 94°C for 2 minutes; 20 cycles of 94°C for 30 seconds, 55°C for 30 seconds, 68°C for 4 minutes; followed by a final extension at 68°C for 10 minutes. All PCR reactions were carried out using Platinum Taq HiFi polymerase. Resulting amplicons were analyzed by gel electrophoresis, purified using the Macherey-Nagel gel and PCR purification kit, and co-transfected with the pSG3Δenv vector (NIH AIDS Reagent Program) into 293T cells to produce pseudoviruses as previously described.

### Neutralization assays

Viruses were tested against broadly neutralizing antibodies (bNAbs) and purified autologous IgGs using the TZM-bl cell neutralization assay, as previously described ^47,84^. All neutralization assays were conducted in laboratories adhering to Good Clinical Laboratory Practice (GCLP) quality assurance standards. Pseudovirus clones derived from both the reservoir and rebound phases were tested. For autologous antibody testing, baseline IgG was purified using Protein G Sepharose 4 Fast Flow (GE Life Sciences). Neutralization assays were performed using a starting maximum concentration of 50 µg ml^-1^ for 3BNC117 and 10-1074, and 500 µg ml^-1^ for autologous IgG, followed by eight five-fold serial dilutions. All experiments were conducted in duplicate. Neutralization breadth was evaluated using a panel of ten HIV-1 Env pseudoviruses selected based on reported resistance to both 3BNC117 and 10-1074 (https://www.hiv.lanl.gov/catnap). The panel included the following Env clones: 700010040.C9.4520, H030.7, H086.8, THRO4156.18, H035.18, H704_1109_140_RE_cs, 191955_A11, 89-F1_2_25, 6322.V4.C1, and 3637.V5.C3.

### Depletion of residual bNAbs from longitudinal IgG samples

To remove residual bNAbs from longitudinal IgG samples, anti-3BNC117 and anti-10-1074 antibodies engineered with a FLAG epitope on the light chain were used as capture reagents. Following IgG purification from plasma, samples were incubated with the FLAG-tagged anti-bNAb-idiotype reagents at a 2-fold molar ratio of the anti-idiotype to the estimated bNAb concentration for 2 hours at 4°C to allow immune complex formation. Samples were then incubated overnight at 4 °C with anti-FLAG M2 magnetic beads (Sigma–Aldrich, cat. no. M8823). Beads were removed magnetically, and the supernatant (depleted of infused bNAbs) was collected, tested for residual bNAbs by ELISA and if below the limit of detection used for downstream autologous neutralization assays. Depletion was repeated if residual bNAb was detected until it no longer was.

### Time-to-event analyses

To evaluate the combined influence of endogenous and therapeutic antibody pressure on time to ART restart, time-to-event analyses were performed using Cox proportional hazards models with time to ART restart as the outcome. Cox proportional hazards analyses incorporated observed ART restart events occurring beyond 96 weeks when available. Baseline reservoir sensitivity to autologous antibodies, 10-1074, and 3BNC117 was quantified using IC80 values, which were log10-transformed and standardized before modeling. Univariable, pairwise (models including two resistance variables simultaneously), and multivariable models were fit to assess the individual and combined contributions of each resistance dimension to rebound timing. Hazard ratios reflect the effect of a one standard deviation increase in resistance. Analyses were performed using Cox proportional hazards regression in GraphPad Prism and R (survival package version 3.8-6).

## Data Availability

All data produced in the present work are contained in the manuscript

## Acknowledgments

We thank all study participants for their time and commitment to this research. We thank the RIO study group ^37^. We are grateful to the Rockefeller University Hospital Research Support Office and nursing staff for their assistance, the members of the Nussenzweig laboratory for helpful discussions, and Masa Jankovic and Tacio Waldetario for expert laboratory support.

## Funding Statement

This work was supported in part by the National Institutes of Health (UM1 AI100663 and R01 AI129795 to M.C. Nussenzweig), REACH Delaney (UM1 AI164565 to M. Caskey), the Einstein Rockefeller-CUNY Center for AIDS Research (1P30 AI124414-01A1), BEAT-HIV Delaney (UM1 AI126620 to M. Caskey), the Bill & Melinda Gates Foundation (INV-008540, INV-002705 and INV-036842), the Stavros Niarchos Foundation through its grant to the SNF Institute for Global Infectious Disease Research at The Rockefeller University, and a grant from Robert Wennett.

M.J. Fumagalli is a Robert Wennett fellow. M.C. Nussenzweig is a Howard Hughes Medical Institute (HHMI) Investigator. This article is subject to HHMI’s Open Access to Publications policy. HHMI laboratory heads have granted a non-exclusive CC BY 4.0 license to the public and a sublicensable license to HHMI for their research articles. Under these licenses, the author-accepted manuscript of this article can be made freely available under a CC BY 4.0 license immediately upon publication.

## Author contributions

M.J.F., M.C. and M.C.N. conceived the study and designed the experiments. J.F., S.F., M.C.N. and M.C. designed the clinical protocols. S.F., J.F. and M.C. coordinated the study, collected clinical data, and recruited participants. T.E., M.L., J.F., S.P., A.C., A.U., O.S., R.S., M.B., S.K., C.O. are clinical investigators responsible for recruitment and participant care. E.F. and L-R. C. conducted trial statistical analysis. P.Z. conducted bNAbs screening for study eligibility. H.B., N.R. and H.B. were responsible for management and operational conduct of the trial. S.C. is the community representative for people living with HIV. M.J.F., A.K., M.A., A.L.R., M.S., B.J.H., and A.G. performed the experiments. C.L., S.M.E. and M.S.S. performed the TZM-bl neutralization assays. M.J.F. and C.B.O. coordinated and processed clinical samples. M.J.F., T.O., M.C. and M.C.N. analyzed and interpreted the data. M.J.F. had full access to the raw data reported in this study. M.J.F., M.J., M.C. and M.C.N. wrote the manuscript with input from all authors. All authors discussed the results, provided critical feedback, and approved the final version of the manuscript.

## Data and code availability

Source data underlying all graphs and summary statistics are provided with this paper. HIV-1 sequence data generated in this study have been deposited in GenBank under accession numbers XXXXX–XXXXX (available after acceptance). No custom computational code central to the conclusions of this study was generated.

De-identified participant-level clinical and laboratory data from the RIO trial, including relevant data dictionaries and supporting study documentation, will be available through a controlled-access process following completion of the trial and database lock, in accordance with participant consent and institutional ethical requirements. Requests for access should be directed to the Chief Investigator and will be reviewed by the Trial Management Group, Independent Data Monitoring Committee, and Trial Steering Committee. Approved investigators will be required to sign a data access agreement before receiving access to the requested datasets. The RIO clinical protocol and statistical analysis plan are available within the clinical manuscript.

## Extended Data Figure

**Extended data Figure 1.**
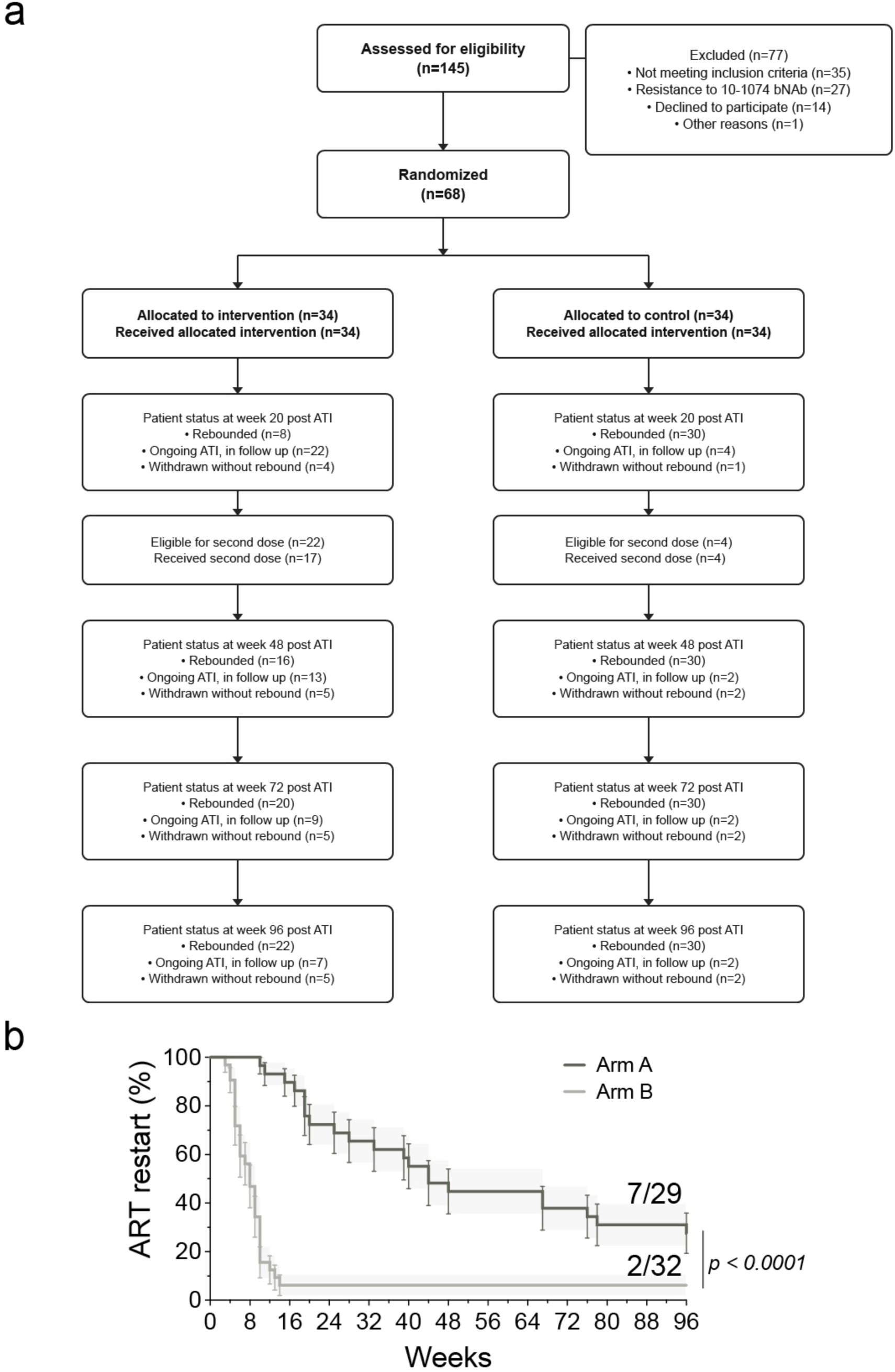
Trial participant selection and time to ART restart after ATI. **a**, CONSORT flow diagram showing participant enrollment, randomization, allocation to Arm A or Arm B, follow-up, ART restart categories, and inclusion in virologic analyses. **b**, Kaplan–Meier curves showing time to ART restart (up to 96 weeks) in participants from Arm A (bNAbs; *n* = 29) and Arm B (placebo; *n* = 32). Curves were compared using the Gehan–Breslow–Wilcoxon test (χ² = 30.09, *P* < 0.0001). Numbers at the end of each curve indicate the number of participants remaining off ART at week 96 relative to the total in each arm.

**Extended data Figure 2.**
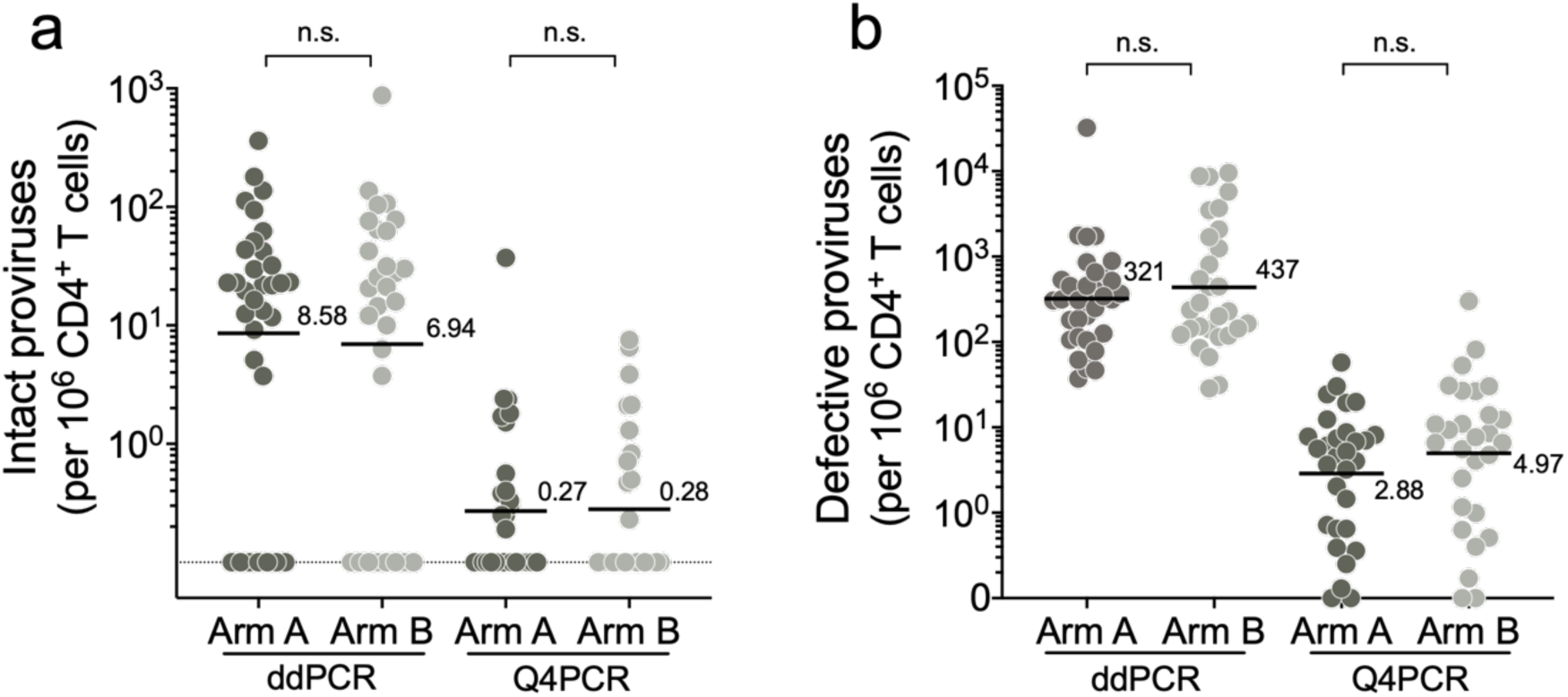
Baseline HIV reservoir in circulating CD4+ T cells measured by ddPCR and Q4PCR. Graphs show the numbers of (**a**) intact and (**b**) defective proviral genomes per million CD4+ T cells. Each dot represents the value for one participant. Arm A participants black and Arm B grey. Sample size: ddPCR – Arm A, *n* = 32; Arm B, *n* = 29; Q4PCR – Arm A, *n* = 30; Arm B, *n* = 27. The black line and numbers indicate the geometric mean. Group comparisons were performed using the Mann–Whitney test, with statistical significance defined as *P* < 0.05.

**Extended data Figure 3.**
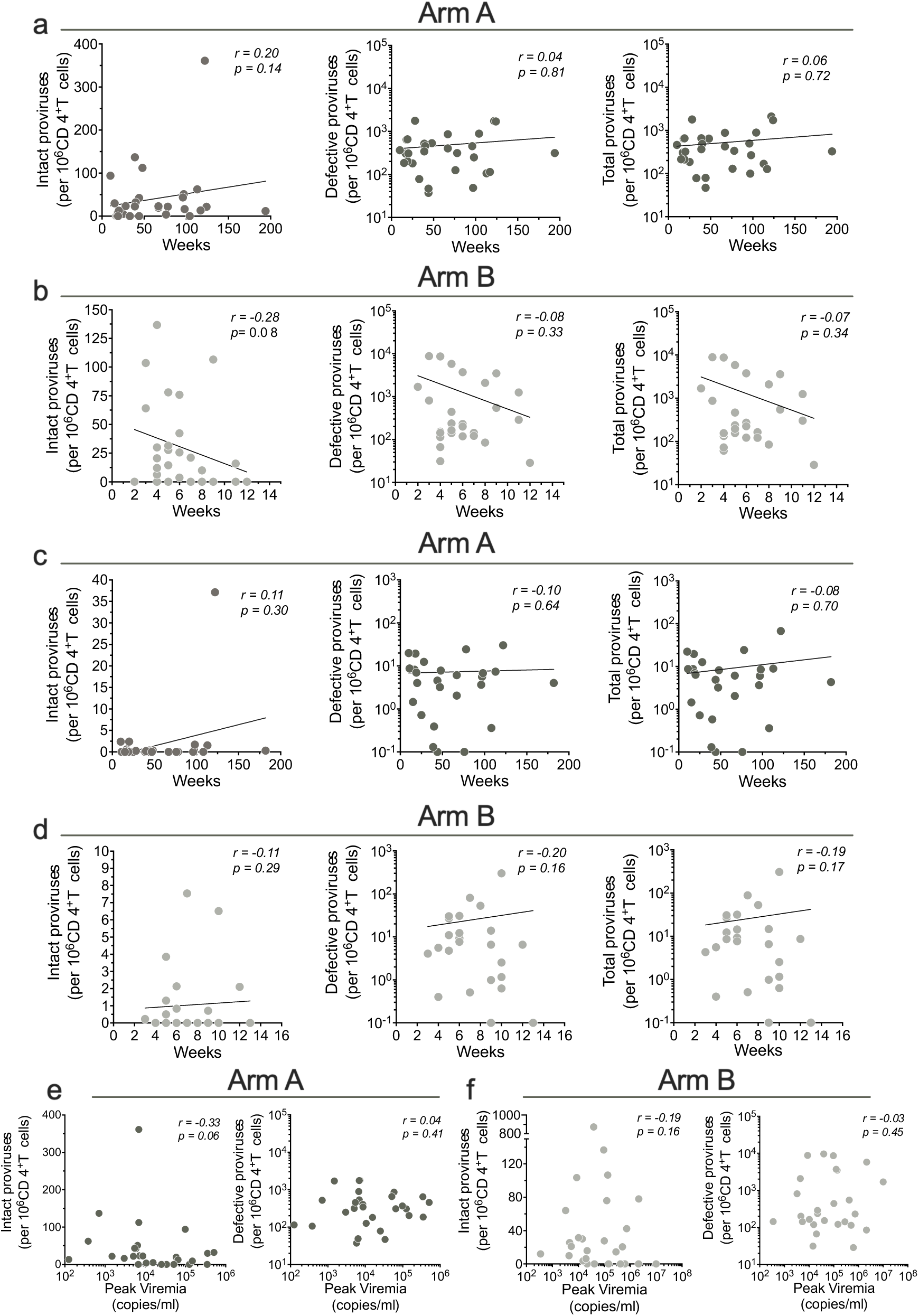
Correlation between baseline intact, defective and total reservoir size, peak viremia and time to ART restart in weeks. (**a - d**) Correlation between baseline intact, defective, and total proviral reservoir size measured by ddPCR (a-b, e-f) or Q4PCR (c,d) and time to ART restart in weeks in Arm A (a,c) and Arm B (b,d). Each dot represents one individual. For ddPCR measurements, n = 29 for Arm A and n = 27 for Arm B. For Q4PCR measurements, n = 26 for Arm A and n = 25 for Arm B. (**e,f**) Relationship between baseline reservoir size measured by ddPCR and peak plasma viremia in Arm A (e) and Arm B (f). Each dot represents one individual. n = 28 for Arm A and n = 27 for Arm B. Y axes indicate the number of intact or defective proviruses, and x axes indicate plasma viremia at peak rebound. Correlation coefficients were calculated using Spearman’s rank correlation with 95% confidence intervals. Nonlinear curve fitting was performed with x as a linear function, and y as a linear or logarithmic function for intact and defective genomes, respectively.

**Extended data Fig. 4.**
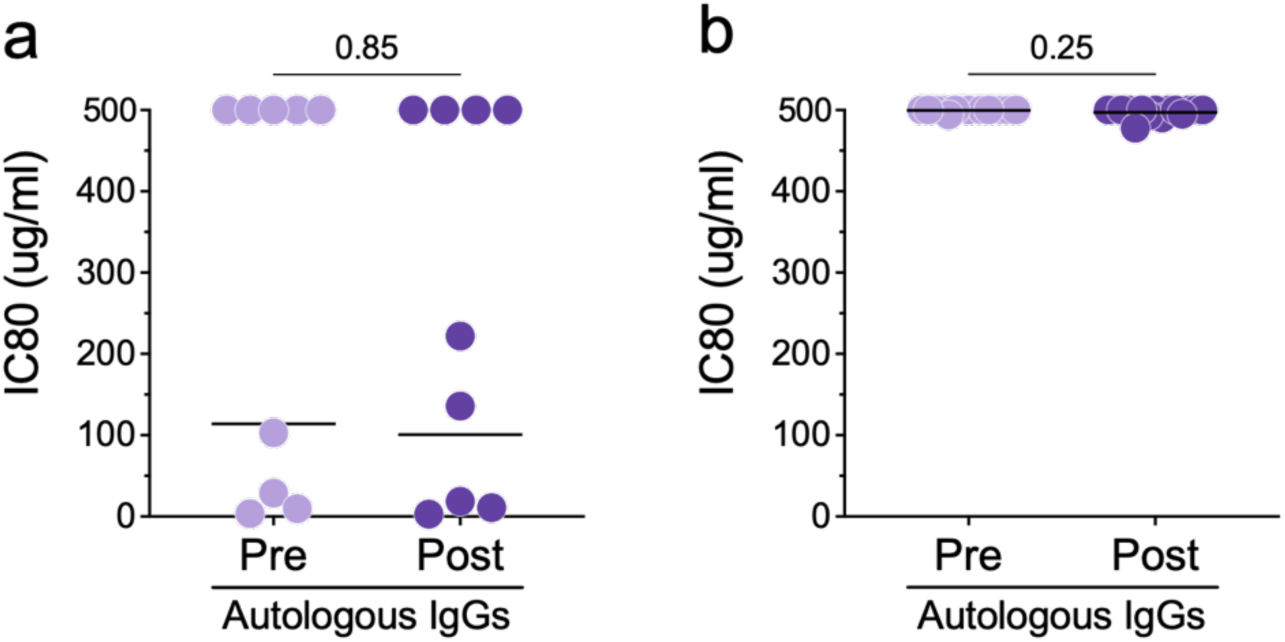
Autologous neutralization activity and breadth post-bNAb therapy in Arm A participants. (**a**) Autologous neutralization sensitivity (IC80) of baseline reservoir-derived pseudoviruses tested against IgG isolated before bNAb infusion and at later post-infusion time points after depletion of residual infused bNAbs. Each circle represents the geometric mean IC80 for an individual participant (n = 9 participants). Post-infusion IgG samples were collected at the following time points, with corresponding residual plasma concentration (µg ml^-1^) of 3BNC117 and 10-1074, respectively: P4, week 39 (12.5 and 10.7); P7, week 64 (4.1 and 4.7); P15, week 134 (<1 and <1); P18, week 64 (13.8 and 18.1); P19, week 36 (25.9 and 30.0); P21, week 119 (2.2 and 3.3); P32, week 123 (<3 and <3); P33, week 27 (338 and 32.5); and P48, week 56 (22.0 and 20.8). (**b**) Neutralization breadth of autologous IgG isolated before and after bNAb therapy was evaluated in a panel of 10 HIV-1 strains resistant to both 3BNC117 and 10-1074. Analyses included 15 participants, with post-infusion IgG samples collected at a mean of 65.7 weeks after infusion. Group comparisons were performed using two-sided Wilcoxon matched-pairs signed-rank test (P < 0.05).

**Extended data Figure 5.**
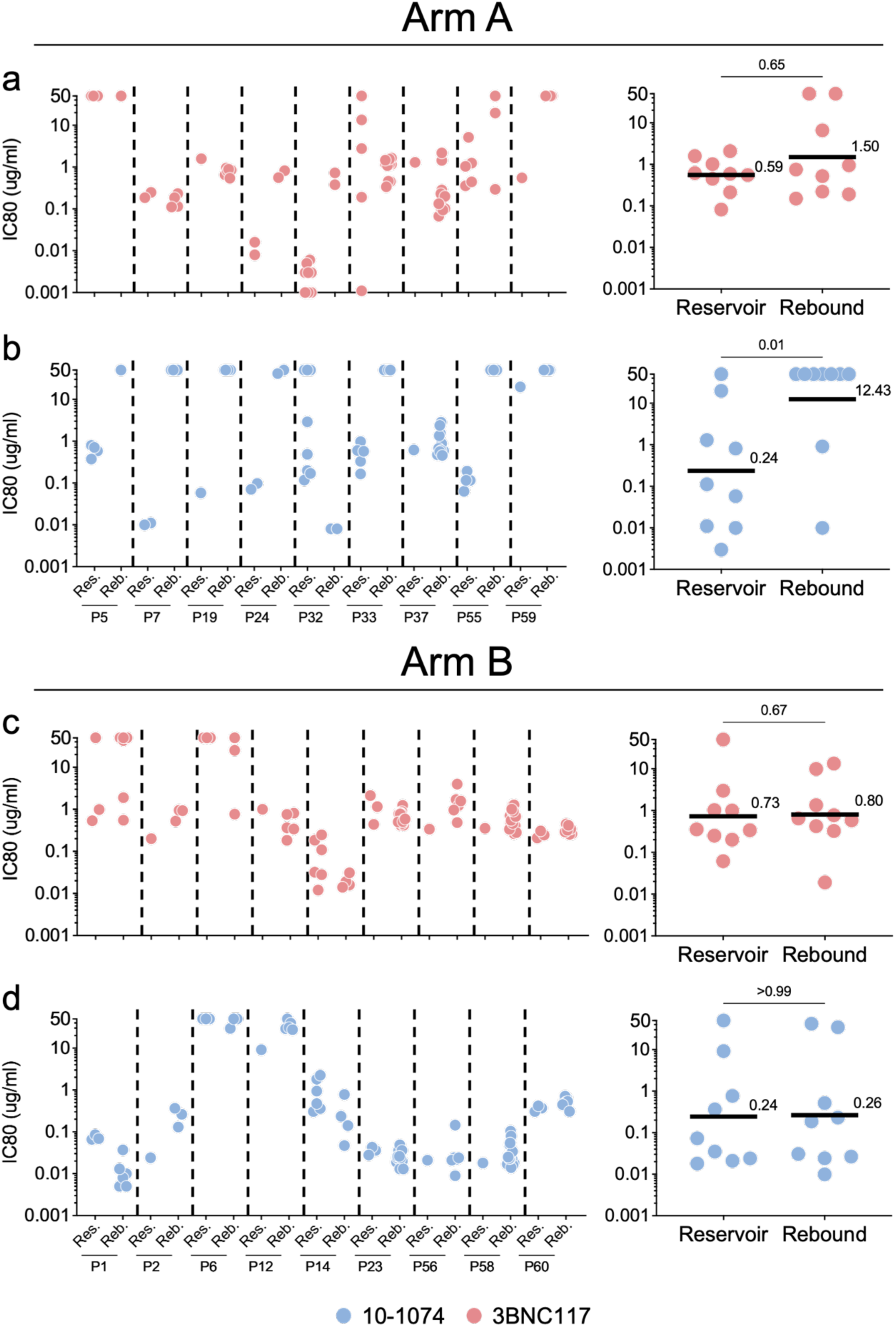
Per-participant matched reservoir- and rebound-derived pseudovirus neutralization sensitivities to 3BNC117 and 10-1074 in both study arms, expressed as IC80 values. **a-d,** Each dot represents a reservoir- or rebound-provirus from each participant in (**a, b**) Arm A and (**c, d**) Arm B. N = 27 participants for each Arm. Members of expanded clones are represented only once irrespective of the size of the clone. Geometric mean IC80 values of all reservoir- and rebound-derived clones per participant are indicated alongside each graph. Group comparisons were performed using the Mann–Whitney test, with statistical significance defined as *P* < 0.05. Black lines indicate the geometric mean.

**Extended data Figure 6.**
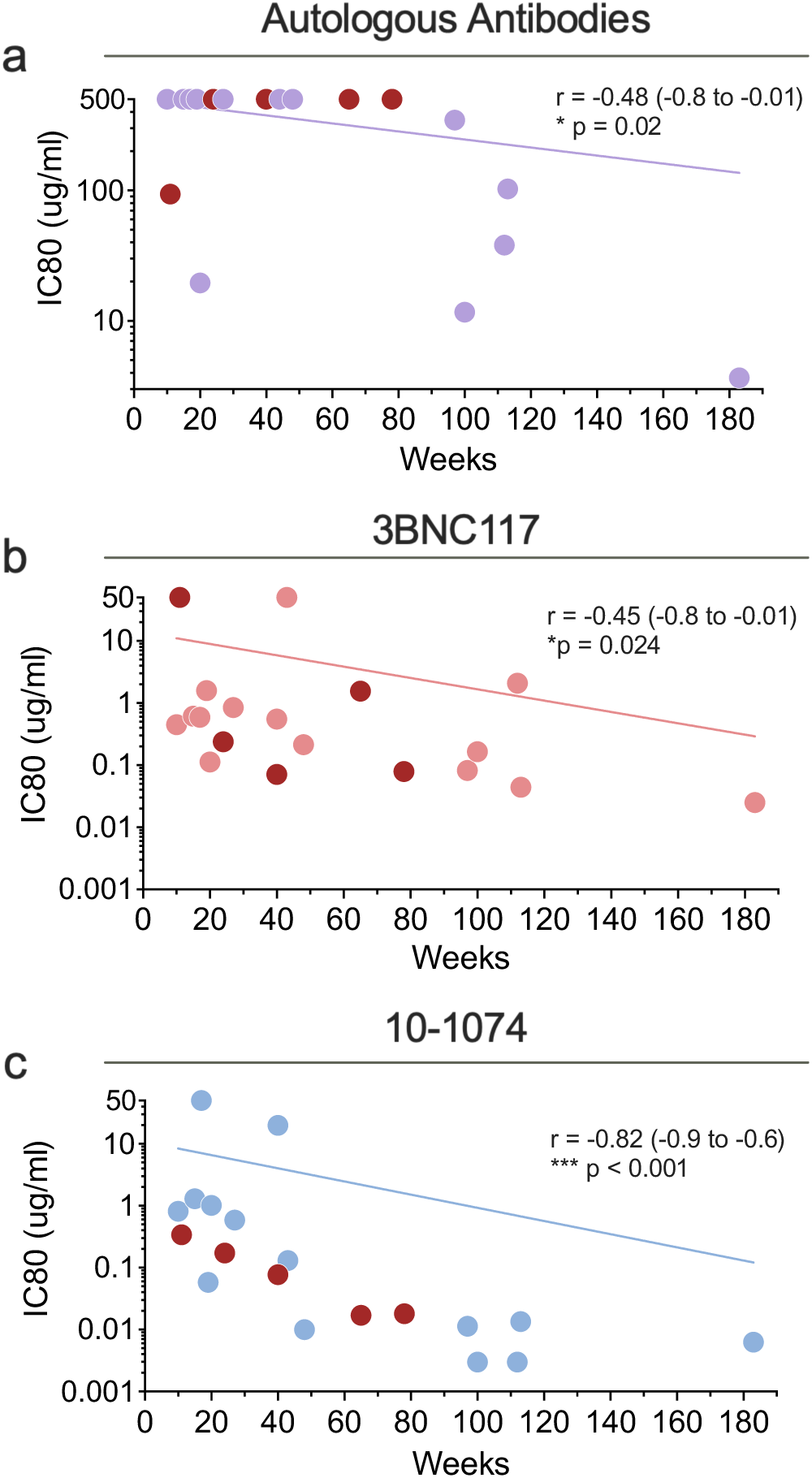
Reservoir sensitivity to autologous antibodies, 3BNC117 and 10-1074 including participants with intact *env* but not confirmed intact proviral genomes. **a-c**, Correlation between time to ART restart and baseline reservoir sensitivity to (**a**) autologous antibodies, (**b**) 3BNC117, and (**c**) 10-1074. Each dot represents the geometric mean IC80 (Y axis) and time to rebound in weeks (X axis) for one participant. Red dots represent participants that we were able to detect intact *envs* but not confirmed intact provirus. Nonlinear curve fitting was performed with *x* as a linear function of time and *y* as a logarithmic function of IC80 values. Correlation coefficients were calculated using Spearman’s *r* with 95% confidence intervals.

**Extended data Figure 7.**
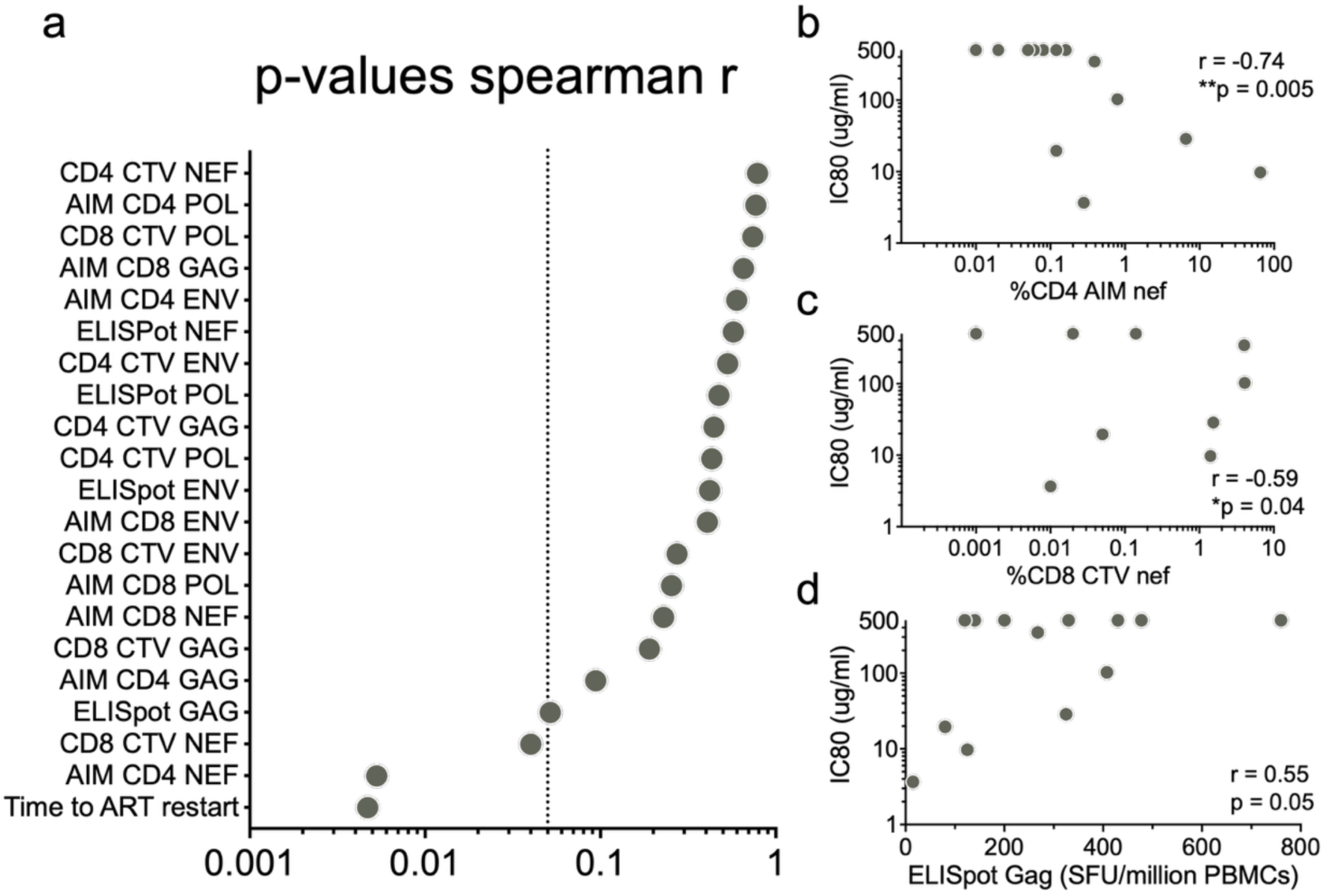
Associations between baseline autologous neutralizing activity and cellular immune responses reported elsewhere^52^. (**a**) Summary of Spearman correlations between baseline autologous neutralizing activity against reservoir viruses and cellular immune responses measured by ELISpot, CTV, and AIM assays at week 0. *P* values are indicated for each assay; the dashed line marks *P* = 0.05. (**b–d**), Scatter plots showing correlations between autologous antibody neutralization (IC₈₀) and (**b**) AIM CD4⁺ responses to Nef, (**c**) CD8⁺ CTV responses to Nef, and (**d**) ELISpot responses to Gag peptides. Each point represents an individual participant.

**Extended Data Figure 8.**
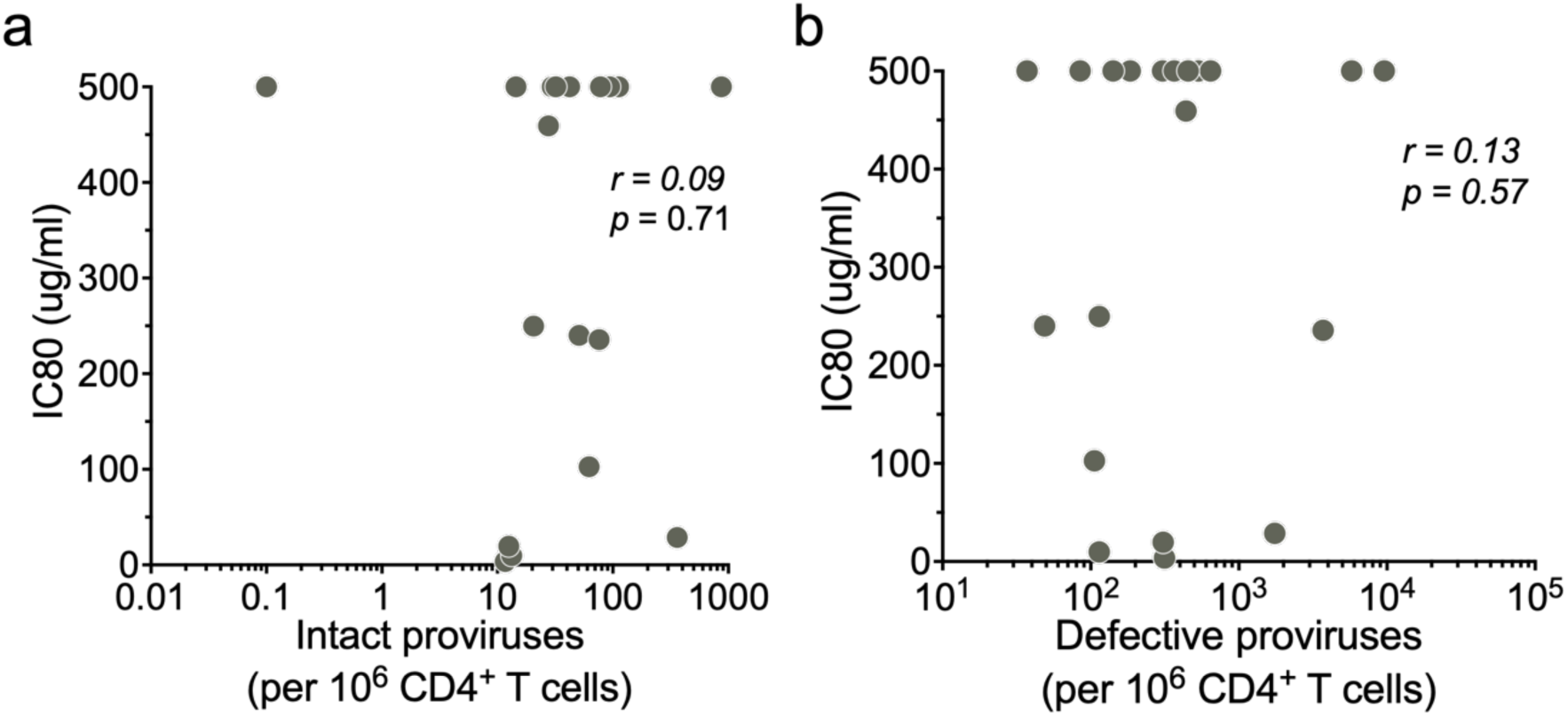
Relationship between baseline reservoir size and autologous neutralizing activity. Baseline (**a**) intact and (**b**) defective proviral DNA levels measured by ddPCR are shown for participants with and without detectable autologous neutralization against reservoir-derived viruses. Correlation coefficients were calculated using Spearman’s *r* with 95% confidence intervals.

**Extended data Fig. 9.**
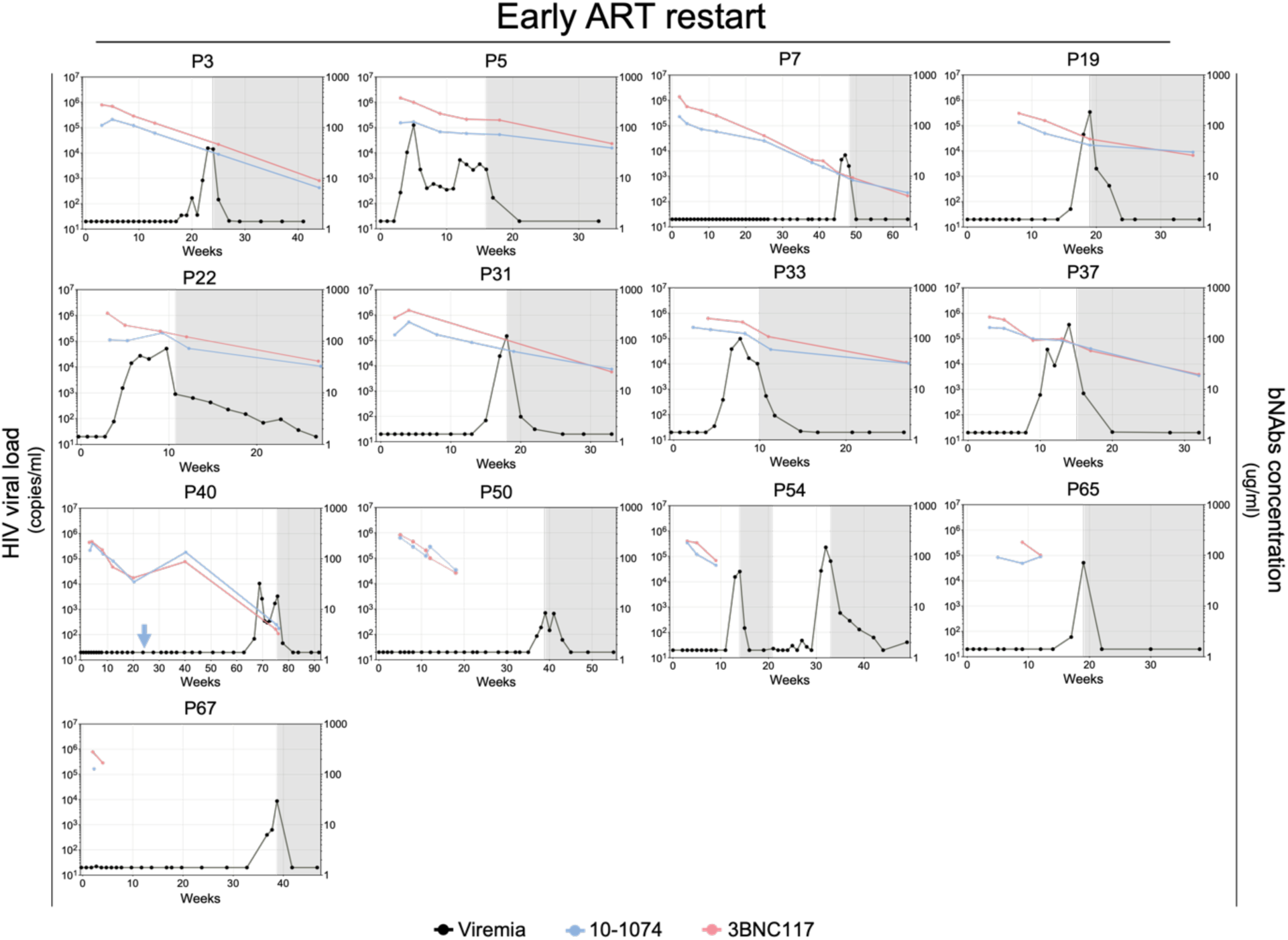
Longitudinal plasma viremia and circulating bNAb concentrations in participants with early ART restart. Longitudinal plasma HIV-1 RNA levels (black) and circulating bNAb concentrations for 10-1074 (blue) and 3BNC117 (red) are shown for participants who restarted ART early, defined by protocol criteria. Time is shown in weeks on the x axis, with HIV-1 RNA copies per milliliter and bNAb concentrations in microgram per milliliter plotted on log10 scales on the y axes. Blue arrows indicate administration of a second bNAb infusion. Gray shading denotes periods on antiretroviral therapy.

**Extended data Figure 10.**
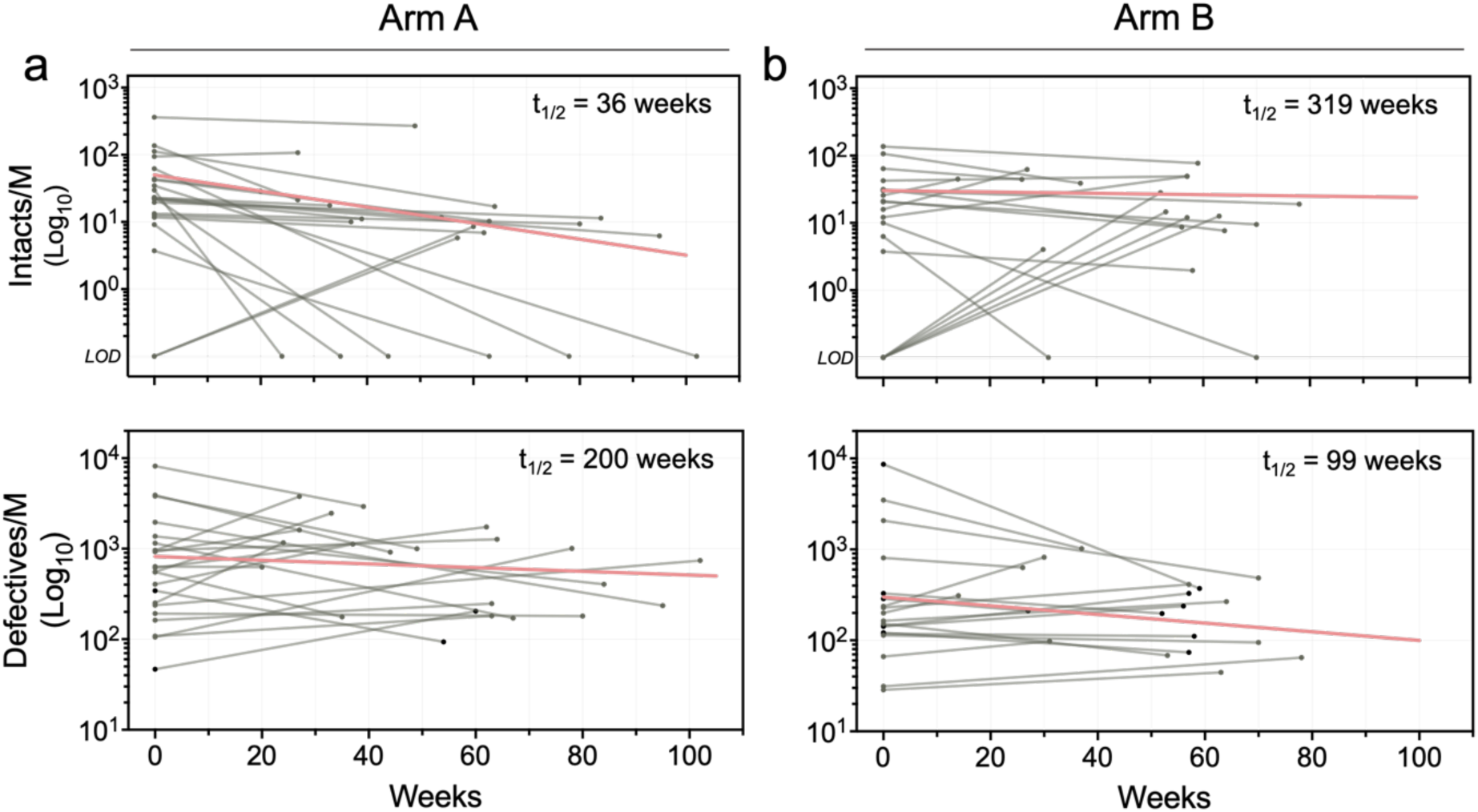
Decay of intact and defective proviral reservoirs measured by ddPCR. (**a,b**) Numbers of intact and defective proviruses quantified by ddPCR at baseline and follow-up time points in participants from Arm A (*n* = 22) and Arm B (*n* = 20). Time in weeks after the first infusion or treatment interruption is shown on the x axis. Each grey line represents an individual participant. Half-life estimates were derived using log-linear regression of log₁₀(*x* + 1)–transformed values; the red line indicates the fitted regression model.

**Extended data Figure 11.**
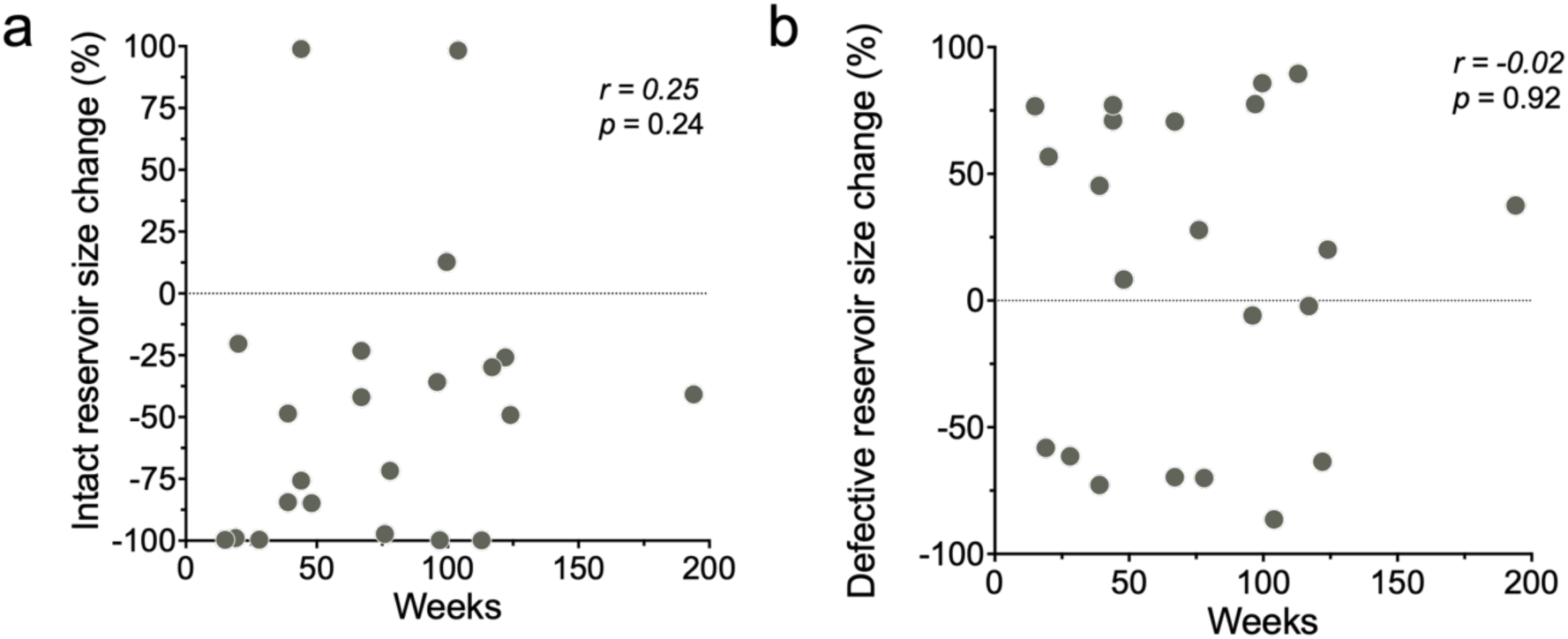
Correlation between the change in reservoir size and time to ART restart. Scatter plots showing the relationship between time to ART restart and the change in (**a**) intact and (**b**) defective reservoir size relative to the pre-infusion baseline of Arm A participants. The x axis indicates time to ART (weeks) and the y axis indicates the percentage change in reservoir size. Each point represents an individual participant. The dotted line indicates 0% change relative to baseline. Correlation coefficients were calculated using Spearman’s *r* with 95% confidence intervals.

## Supplementary Tables (provided as separate excel files)

**Supplementary Table 1.** Baseline demographic and clinical characteristics of RIO participants.

**Supplementary Table 2.** Quantification of HIV reservoir size by digital droplet PCR (ddPCR) and quantitative quadruplex four-probe PCR (Q4PCR) for individual participants.

**Supplementary Table 3.** Neutralization sensitivities and baseline near full-length proviral sequences from participants in Arms A and B.

**Supplementary Table 4.** Neutralization sensitivities and individual ENV coding region sequences obtained from plasma of participants in Arm A and B during rebounding viremia.

**Supplementary Table 5.** Geometric mean neutralization sensitivities of reservoir-derived and rebound pseudoviruses from individual participants in both study arms, tested against baseline autologous IgG, 3BNC117, and 10-1074.

**Supplementary Table 6**. Serum concentrations of 3BNC117-LS and 10-1074-LS in participants with prolonged viral control.

**Supplementary Table 7**. Cox proportional hazards models evaluating the relationship between time to viral rebound and baseline reservoir sensitivity to autologous antibodies, 10-1074, and 3BNC117.

